# Large language models enable consensus-level interpretation in metagenomic diagnostics

**DOI:** 10.64898/2026.07.29.26358751

**Authors:** Eike Steinig, Marcelina Krysiak, Kirti Deo, Andrew Duncan, Jacqueline Prestedge, Jeremy Barr, Jean Moselen, Sadid F. Khan, Janath A. Fernando, Ivana Savic, Bhargavi Yellapu, Ammar Aziz, Wytamma Wirth, Jessica Parry, Angela McDonald, Chhay Lim, Sharon Trevor, Benjamin Aw-Yeong, Georgia McCluskey, Michael Moso, Eddie Chan, Sonia L. La Vita, Penelope A. Bryant, Amy Crowe, Ramla Maalim, Diana Velasquez Reyes, Maryza Graham, Eloise Williams, Jason C. Kwong, Rachel Woolstencroft, Monica Slavin, Lyndell L Lim, Lachlan J.M. Coin, Leon Caly, Katherine Bond, Chuan Kok Lim, Timothy P. Stinear, Deborah A. Williamson, Prashanth S. Ramachandran

## Abstract

Metagenomic sequencing can detect a broad range of pathogens, but interpreting which detections are clinically relevant requires expert adjudication that is difficult to scale and standardize. Here we present diagnostic classifiers that formalize expert adjudication by combining structured decision trees with large language model reasoning to assign diagnoses and select pathogen candidates. We first developed a short-read metagenomic assay for sterile-site specimens (cerebrospinal and ocular fluid) in the META-GP study (Victoria, Australia, 2024-2025) and evaluated classifiers on a validation dataset (n = 96; clinical samples, spike-ins and controls). Locally deployed, open-weight reasoning models (Qwen3) achieved diagnostic performance comparable to expert consensus, improving with clinical context (n = 79, above experimental limit-of-detection; without clinical notes, 94.4% sensitivity, 95.4% specificity; with clinical notes, 97.2% sensitivity, 100% specificity). Automated adjudication enabled systematic benchmarking of computational parameters and regression testing for pathogen detection tasks. In a heterogeneous development cohort (n = 78), reviewers and classifiers identified clinically significant pathogens missed during routine testing. By reproducing consensus detections without requiring a full review panel, diagnostic classifiers enable scalable, standardized metagenomic interpretation that complements expert adjudications.

## Introduction

Metagenomic sequencing has emerged as a powerful tool for diagnostics, offering the ability to detect a broad range of pathogens without prior assumptions about the cause of disease. However, the interpretation of metagenomic results depends heavily on expert adjudication (Wilson2019, Chiu2019, Miller2019, Blauwkamp2019, Gu2020, Ramachandran2022, Fourgeaud2024, Benoit2024, Alcolea-Medina 2025). In practice, clinicians and laboratory specialists must determine whether detected organisms represent true pathogens, background commensals, environmental contaminants or incidental findings, which requires contextual judgment beyond quantitative thresholds.

This challenge arises because untargeted metagenomic sequencing inevitably captures nucleic acid from non-pathogenic microorganisms present in clinical samples or introduced during sample processing. As a result, organisms with no causal relationship to disease may exceed reporting thresholds, while clinically relevant pathogens may be detected below thresholds (Benoit2024, Torres-Montaguth 2025). Determining clinical relevance therefore requires synthesizing multiple sources of information, including sample type, organism identity, relative abundance, contamination risk, patient clinical history, and results from orthogonal diagnostic tests (Gu2020, Ramachandran2022, Alcolea-Medina2025, Blauwkamp2019, Fourgeaud2024, Benoit 2024). However, this multidimensional reasoning is difficult to formalize within current analytical and interpretation frameworks (Meyer 2022).

In operational diagnostics, the reliance on expert review has become a major bottleneck in both the evaluation and clinical deployment of metagenomic diagnostic assays (**Appendix**). Expert adjudication is time- and labour-intensive and requires sustained access to specialized multidisciplinary expertise, often unavailable outside tertiary centres. Even minor changes to laboratory protocols or computational pipelines typically necessitate a complete re-adjudication of a reference dataset for quality assurance. This constrains systematic parameter optimization and renders regression testing impractical for robust quality assurance. Cognitive biases, such as anchoring on familiar taxa or recalling prior reference cases, also undermine consistency and reproducibility across reviewers and institutions. These limitations highlight the need for an interpretive framework that can capture the contextual reasoning embedded in expert adjudication. Rather than relying on fixed abundance thresholds, such a framework would assess whether detected organisms are plausible causes of disease given the clinical context of each patient.

Recent advances in large language models (LLMs) suggest a path towards this goal. By integrating structured metagenomic outputs with unstructured clinical information, LLMs offer a means to formalize clinician-like reasoning over complex diagnostic evidence. Instead of asking whether a pathogen exceeds a predefined quantitative threshold, LLM-based systems could evaluate whether its detection is consistent with known disease presentations, patient history, and supporting diagnostic data. Advances in medical foundation models (Liu2025, McDuff 2025) and open-weight LLMs (Sandmann2025, Tordjman 2025) further position this approach as clinically translatable and align with regulatory requirements for diagnostic testing.

In this study, we aimed to develop and validate a metagenomic diagnostic assay for low-biomass sample types that explicitly formalizes expert adjudication. We introduce a tiered ensemble filtering strategy that replaces static abundance thresholds with confidence- and context-aware prioritization of taxonomic detections, enabling systematic interpretation of both above- and sub-threshold findings. Building on this foundation, we design and validate diagnostic classifiers that combine open-weight LLMs with decision trees and patient clinical context to emulate expert adjudication and automate pathogen candidate selection.

## Results

### Analytical validation of the metagenomic assay

We first assessed analytical sensitivity of the assay in a limit of detection (LOD) experiment using multiple taxonomic identification strategies at species rank (**Figure 1**). K-mer and alignment classifiers demonstrated high sensitivity without quantitative abundance thresholds (LOD_95_ = Ct 35.1-37.4 ≈ 28-196 copies/mL); application of common thresholds for short-read metagenomic assays (10 RPM or RPM-ratio) (Miller2019, DeVries2021, Benoit 2024) caused expected reductions in sensitivity across organisms (LOD95 = Ct 29.1 - 35.4 ≈ 602-28322 copies/mL) (**Figures S1-S2**). We next evaluated diagnostic performance of the assay in a validation cohort of patients with suspected central nervous system (CNS) infections in Victoria (Australia, 2024-2025) (n = 96, comprising 87 orthogonally verified clinical and spike-in samples across CSF and ocular fluids; 43 negative, 44 positive, and 9 controls, **Figure S3-S4, Table S1-S2**). Eight positive clinical samples had orthogonal testing results below the experimental LOD_95_ (PCR where available); of these, four DNA and RNA viruses (HHV-6, HIV-1, HSV-2, HTLV-1) could not be detected with any classifier (**Table S3**).

**Figure 1.**
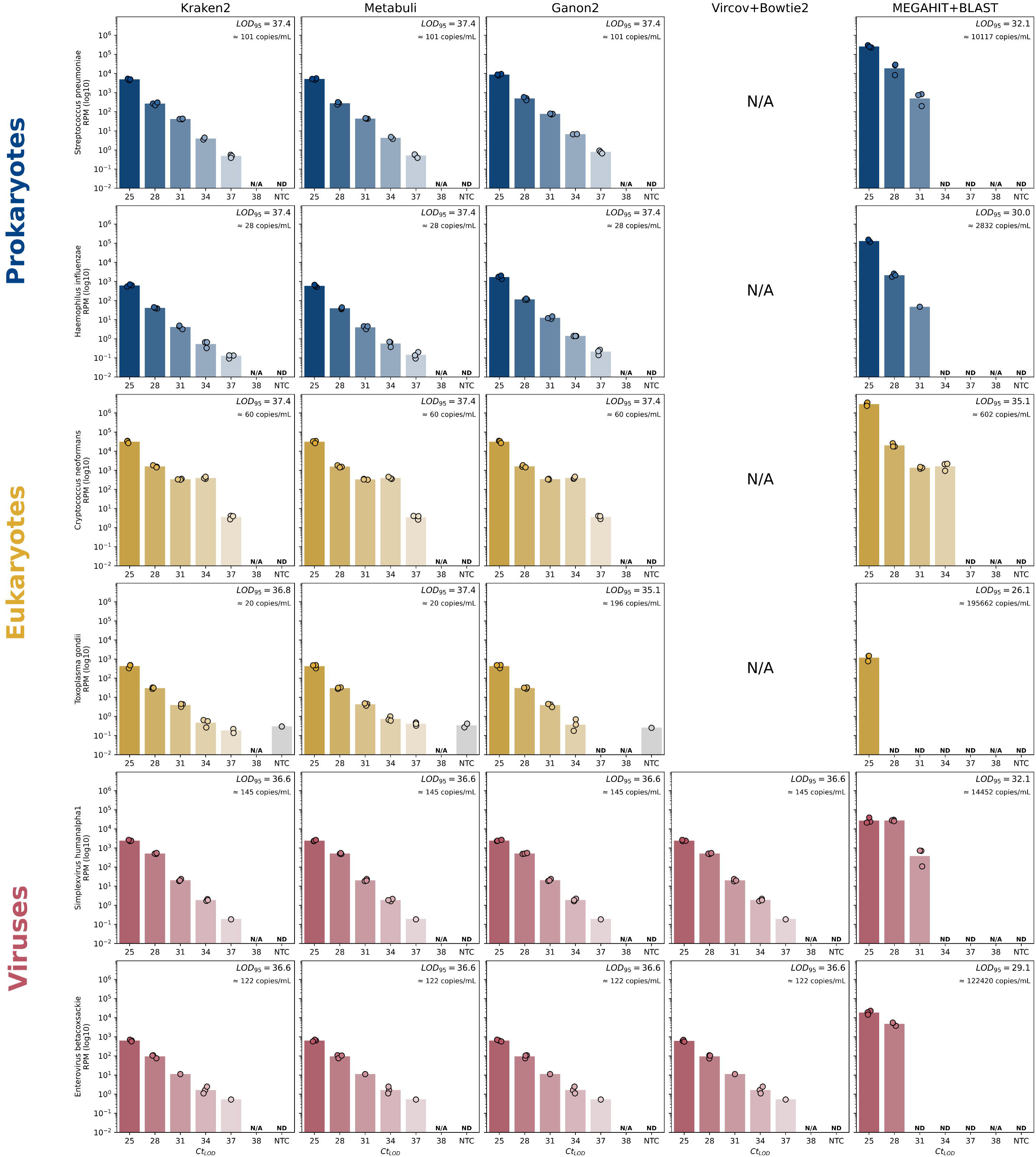
Limit of detection experiments and taxonomic classifier benchmarks for a short-read metagenomic sequencing assay. Limit of detection (LOD) of representative central-nervous system pathogens (rows) spanning prokaryotes (*Streptococcus pneumoniae, Haemophilus influenzae*), eukaryotes (*Cryptococcus neoformans, Toxoplasma gondii*) and viruses (*Simplexvirus humanalpha1, Enterovirus betacoxsackie*) assessed at species rank over a dilution series (Ct 25-37) for k-mer read classifiers, viral read alignment and metagenome assembly (columns). Bars, sequence abundance (RPM, log₁₀) or total assembled base-pair abundance (BPM, log₁₀); points, library replicates; annotations, LOD₉₅ (nominal probit) and approximate concentration (copies/mL). RPM, reads per million; BPM, bases per million (metagenome assembly; scaled x 1000 for visual comparability in display only, not for probit analysis); N/A, not applicable to domain; ND, not detected; NTC, no-template control.

During assay development with Cerebro v0.10.0, a review board of laboratory and clinical specialists (PR, AD, MK, KD, ES) interpreting the unstructured, abundance-ranked results from ensemble classifiers achieved 94.4% sensitivity and 97.7% specificity (n = 79 samples above experimental LOD; 77.8% sensitivity and 97.6% specificity including samples below LOD, n = 87) but made three characteristic errors in clinical samples: (1) an orthogonally verified low-abundance *Simplexvirus humanalpha2* (HSV-2) detection was missed; (2) an orthogonally verified low-abundance *Cryptococcus gattii* detection was missed; (3) a background organism (*Lactococcus lactis*) was selected as a candidate pathogen in an orthogonally negative ocular fluid sample (**Table S1**). These two failure modes (overlooking true positives that fall below reporting thresholds, and elevating contaminant or commensal organisms) occurred despite the board’s familiarity with the clinical samples and the composition of the validation dataset, making these adjudication results an optimistic estimate. This stimulated the development of a systematic stratification framework to (computationally) formalize the above- and sub-threshold adjudication process described by Benoit and colleagues (Benoit 2024).

### Tiered stratification establishes diagnostic performance ceiling

We used negative control comparisons, prevalence contamination filters, domain-specific abundance thresholds and ensemble classifier congruence to assign taxa to primary- (above-threshold, high-abundance, high-confidence), secondary- (below-threshold, low abundance, medium-high confidence) and target-tiers (high-priority organisms, low-abundance) (**Figure 2, Methods**). This approach controls for divergent genome characteristics across taxonomic domains and for variable performance of individual classifiers in taxonomic identification (McLaren2019, Ye2019, Meyer 2022). Applying a configuration empirically optimized for sensitivity (using the LOD experiment data) caused a single verified positive (also missed by the review board) to fall below thresholds (C. *gattii*, 4 reads with Metabuli). The performance ceiling for the validation dataset using the stratification filter in this configuration was therefore 97.2% sensitivity and 100.0% specificity (n = 79, samples above experimental LOD). Configuration of global abundance thresholds across tiers without other filter-controls (≥ 10 RPM primary, 1-10 RPM secondary, 0-1 RPM target) demonstrated that full ensemble filtering markedly reduced the number of taxa called per sample across domains and reporting tiers, even in the sensitivity configuration (**Figure 2**).

**Figure 2.**
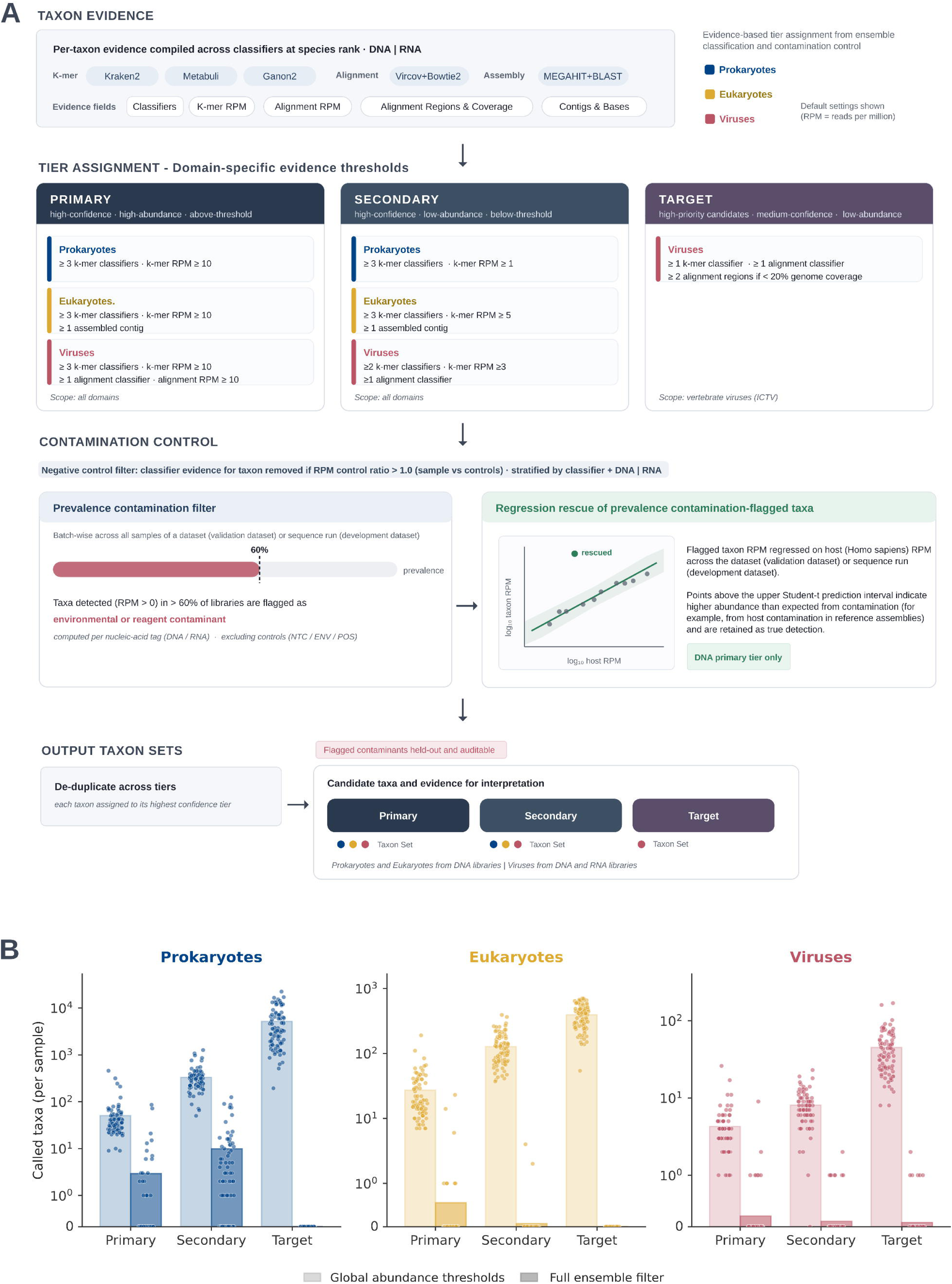
Tiered filtering scheme for ensemble classification and interpretation of taxonomic identifications. (A) Stratification framework assigning species-rank detections to primary (above-threshold), secondary (below-threshold) and target (curated high-priority) tiers using negative-control comparisons, prevalence contamination filtering with outlier rescue, domain-specific abundance thresholds and ensemble classifier congruence. (B) Number of called taxa per sample (log₁₀) by taxonomic domain (columns) and reporting tier (primary, secondary, target), comparing global abundance thresholds (≥10 RPM primary, 1-10 RPM secondary, 0-1 RPM target) (light bars) with the full ensemble filter (dark bars). Bars, mean; points, individual samples. RPM, reads per million; NTC, no-template negative control; ENV, environmental negative control; POS, positive sequence run control.

### Consensus adjudication mitigates individual reviewer variability

We independently assessed diagnostic performance of the stratified results (the same results available to the review board) with a group of clinical microbiologists who were blinded from the clinical cases and composition of the validation dataset. We intentionally evaluated untrained reviewers without clinical notes to establish a conservative baseline of standalone specialist performance, isolating the interpretive burden from prior familiarity or contextual anchoring. This design reflects a worst-case scenario for individual adjudication but does not represent the performance ceiling of experienced mNGS interpreters working with full clinical context. Clinical microbiologists adjudicated the stratified results through the Cerebro interface (v0.12.0) without task-specific training or clinical notes (n = 10, **Methods**). Individual adjudications from untrained reviewers were variable, yielding 77.9% average sensitivity (± 15.0 SD; 95% CI: 67.2-88.6%; range: 58.3-97.2%) and 97.3% average specificity (± 3.2 SD; 95% CI: 95.0-99.6%; range: 89.1-100%) (**Figure 3A-C**). A single reviewer (R10) reached the performance ceiling without errors; in total, four reviewers achieved greater than 80% sensitivity and specificity (**Figure 3A**). Consensus across all reviewers mitigated this variability and came close optimal performance (97.2% sensitivity, 97.7% specificity, **Figure 3C**). The reliability of expert interpretation therefore depended on aggregating multiple independent reviews.

**Figure 3.**
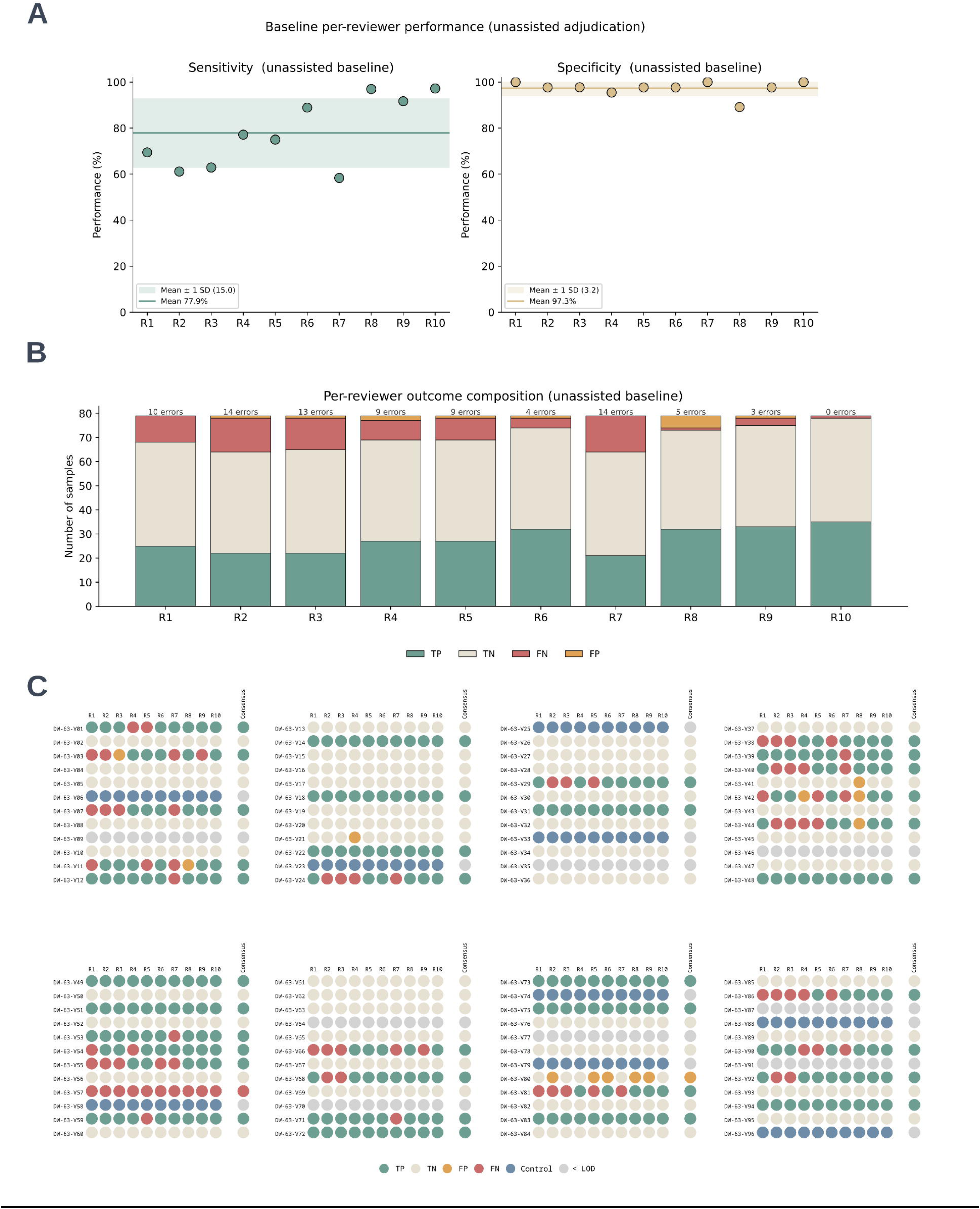
Baseline evaluation of diagnostic performance for pathogen detection by clinical microbiologists. Diagnostic performance of individual clinical microbiologists (n = 10) adjudicating tiered results without task-specific training or clinical notes. (A-C) Per-reviewer sensitivity and specificity, outcome composition, and reviewer performance, including consensus calls, against the orthogonal reference tests. One consistent false negative call from a threshold filtered *Cryptococcus gattii* (DW-63-V57) was not possible to select and is included in the outcome composition plot (B) but not in the error text above each bar. TP, true positive; TN, true negative; FP, false positive; FN, false negative; LOD, limit of detection; SD, standard deviation.

### Diagnostic classifiers match expert performance

Having established multi-reviewer consensus as the performance benchmark, we next designed diagnostic classifiers to emulate expert adjudication. Diagnostic classifiers require no task-specific training and interpret above- and sub-threshold species detections along configurable decision trees using LLMs of the Qwen3 series (QwenAI, Yang 2025) with quantization and reduced parameters suitable for inference on consumer-grade GPUs (META-GPT; **Methods**, **Figure S5-S6, Table S4-S5,** GGUF files and quantization by <u>UnslothAI</u>). At each node of the decision tree, models were prompted to classify a sample as infectious or non-infectious and, for samples deemed infectious, to select a single pathogen candidate. Diagnostic performance was evaluated across model parameters (**Figure 4**) using the same filtered validation results provided to expert reviewers (without clinical notes) so that human and model interpretation were assessed on the identical inputs and context.

**Figure 4.**
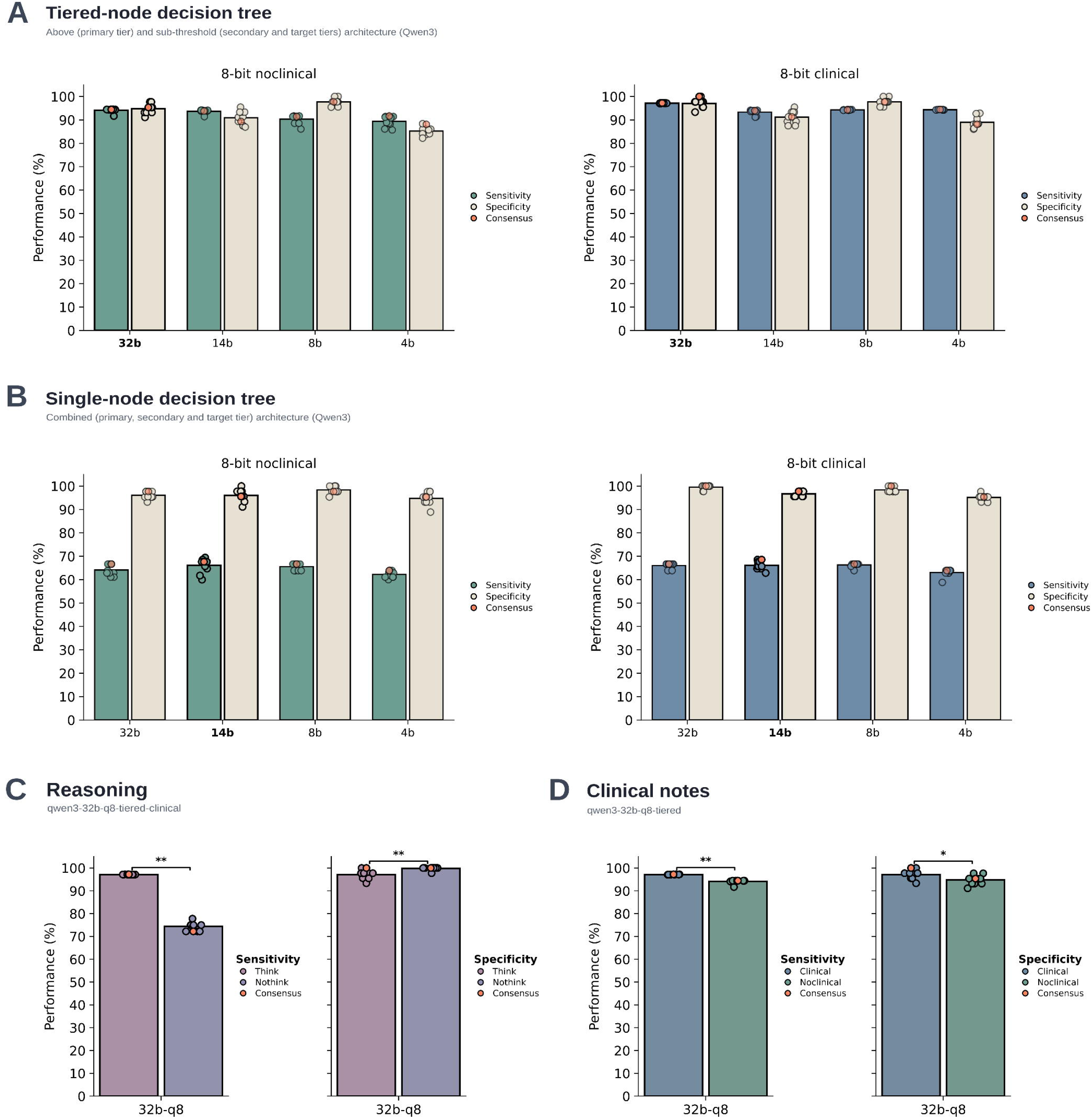
Validation of diagnostic classifier performance and architectures. Diagnostic performance of META-GPT classifiers (Qwen3) across model parameters and decision-tree architectures, evaluated on the tiered validation dataset results. (A-B) Per-replicate and consensus (orange) sensitivity and specificity, with (blue) and without (green) clinical notes, evaluated against orthogonal reference testing for tiered- and single-node decision trees. Best performing models highlighted with bolded labels, data points and bar outlines. (C) Effect of reasoning on diagnostic performance for the best performing model (qwen3-32b-q8-tiered-clinical). (D) Effect of clinical context on diagnostic performance in the best performing architecture (qwen3-32b-q8-tiered). Model identifiers denote parameter count, quantization, decision tree architecture and inclusion of clinical notes (qwen3-32b-q8-tiered-clinical: 32 billion parameters, 8-bit quantization, tiered decision tree with clinical notes). Wilcoxon signed-rank test: **, p < 0.01; *, p < 0.05; TP, true positive; TN, true negative; FP, false positive; FN, false negative; 4b/8b/14b/32b, model parameters (billions); q8, 8-bit quantization.

In the tiered decision-tree architecture without clinical notes, pathogen detection of the best performing model (*qwen3-32b-q8-tiered-noclinical*) across 10 replicates yielded 94.1% average sensitivity (± 0.86 SD; 95% CI: 93.5-94.7%; range: 91.7-94.4%) and 94.7% average specificity (± 2.1 SD; 95% CI: 93.3-96.2%; range: 91.1-97.7%) (**Figure 4A**, green). Consensus across classifier replicates marginally improved the classifier to 94.4% sensitivity and 95.4% specificity (**Figure 4A**, orange data point). The classifier’s lower consensus performance was caused by diagnostic errors in three clinical samples: two above-threshold false positives driven by commensal skin organisms in negative CSF (*Staphylococcus epidermidis*, DW-63-V15; *Staphylococcus capitis*, DW-63-V41) and one sub-threshold false negative in a positive clinical CSF (*Haemophilus influenzae*, DW-63-V38). Overall, the classifier (using a single model with non-zero temperatures) produced more consistent errors among replicates than the human panel (**Figure S7**). Tiered decision-tree architectures and chain-of-thought reasoning (“thinking” mode in Qwen3) were required to maintain sensitivity (**Figure 4B-C**).

We next assessed the effect of clinical context by providing the diagnostic classifiers with short clinical notes where available (48/74 individual patients; 14 positive, 34 negative) together with synthetic notes for spike-in samples (10/10; **Methods**, **Table S1**). With clinical context, the best-performing model (*qwen3-32b-q8-tiered-clinical*) improved to 97.2% sensitivity and 100% specificity from its non-contextualized consensus, reaching the performance ceiling (**Figure 4A**, blue). Average sensitivity (97.1% ± 0.05 SD) and specificity (97.1% ± 1.8 SD) were significantly higher with clinical notes than without (Wilcoxon signed-rank test, sensitivity: p = 0.002; specificity: p = 0.039, **Figure 4D**). All three discordant cases favoured the contextualized classifier, with no regressions (mid-p McNemar’s test, b = 3, c = 0, p = 0.125) with only three discordant pairs the comparison cannot reach significance even when every case favour one method, so this is reported descriptively. Each corrected case involved a commensal or low-abundance organism causing the errors in the non-clinical evaluation (also a source of some uncertainty during expert adjudication, **Figure S7**). Clinical context in the classifiers therefore acted principally to disambiguate plausible-but-spurious detections. Notably, multi-reviewer consensus (n = 10) resolved these cases correctly even without access to clinical notes (**Figure 3C**).

### Expert oversight of a high-performing classifier

To assess model-assisted adjudication, reviewers who had completed the unassisted baseline (n = 8 of 10) repeated the interpretation of the validation cohort after an interval of approximately six months, this time performing a correction task in which the classifier’s proposed diagnosis (*qwen3-32b-q8-tiered-clinical*) was pre-populated in the interface for endorsement or override, with clinical notes provided to both reviewers and model (**Methods**). Because reviewers were anchored on the classifier’s calls, had access to clinical notes withheld at baseline, and the task differed in structure from the unassisted baseline (**Methods**) we report this exercise descriptively as a measure of expert oversight over the classifier’s outputs rather than as a statistically comparable measure of performance improvement (**Figure 5A-C**). Since the classifier reached the performance ceiling on this dataset (97.2% sensitivity and 100% specificity) the exercise assessed whether expert oversight preserved or degraded a strong classifier instead of correcting a weak one.

**Figure 5.**
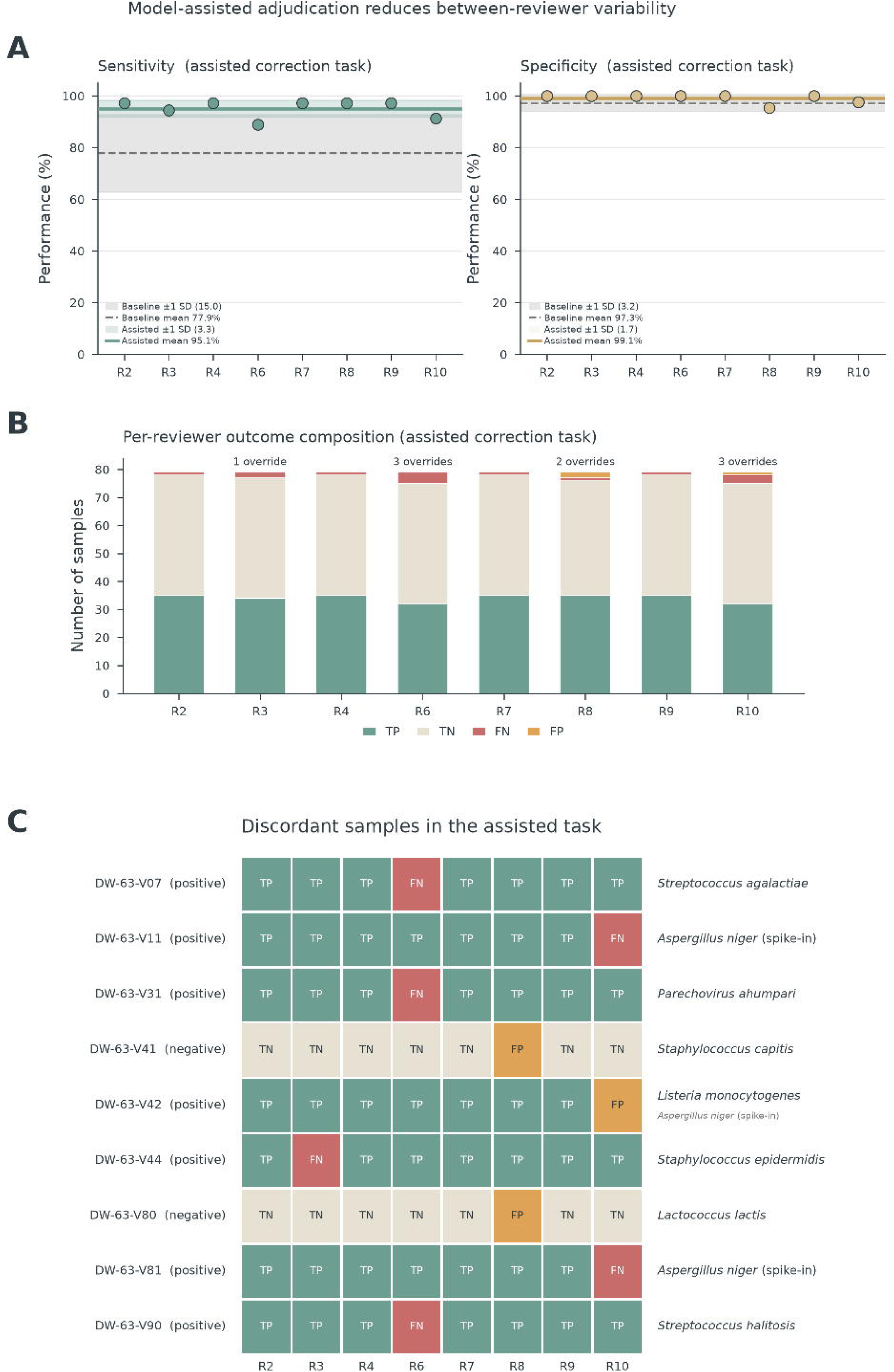
Large language model-assisted evaluation of pathogen detection by clinical microbiologists. Model-assisted correction task in which reviewers (n = 8) endorsed or overrode pre-populated classifier calls (qwen3-32b-q8-tiered-clinical), with clinical notes provided to reviewers and model. (A-C) Per-reviewer sensitivity and specificity relative to the unassisted baseline, endorsement and override outcomes, and reviewer-by-sample outcome matrix with species annotations denoting identity of erroneous overrides (smaller font denotes true positive for spike-in DW-63-V42). One consistent false negative call from a threshold filtered *Cryptococcus gattii* (DW-63-V57) was not possible to select, it is included in every bar in the outcome composition plot (B) but excluded from the override counts above each bar, and is omitted from the discordant-sample matrix (C), which shows only samples on which reviewers disagreed. Model identifiers denote parameter count, quantization, decision tree architecture and inclusion of clinical notes (qwen3-32b-q8-tiered-clinical: 32 billion parameters, 8-bit quantization, tiered decision tree with clinical notes). TP, true positive; TN, true negative; FP, false positive; FN, false negative; SD, standard deviation.

Model-assisted adjudication markedly reduced between-reviewer variability relative to the unassisted baseline, raising individual performance close to the ceiling that unassisted reviewers reached only by consensus (**Figure 5A**, **Figure 3**). Individual sensitivity increased from a mean of 77.9% sensitivity (± 15.0 SD) at baseline to 95.1% (± 3.3 SD), and specificity from a mean of 97.3% (± 3.2 SD) to 99.1% (± 1.7 SD). Reviewers endorsed the classifier’s proposed call in 98.6% of assessments (615/624, n = 78, excluding false negative calls from the filtered *C. gattii* as it presented no selectable call) and the consensus of the assisted adjudications reproduced the classifier’s own performance on the full dataset (97.2% sensitivity, 100% specificity). The nine overrides (1.4% of assessments) each moved away from a correct call (**Figure 5B-C**). Six reversed a correct positive to a false negative, and three introduced a false positive (two negatives reclassified as positive, and one positive assigned an incorrect pathogen). Notably, two reviewers (R6 and R10) who had performed well in the baseline round accounted for most of the incorrect overrides (**Figure 5C**). Four of eight reviewers made no errors in the model-assisted adjudication, and were in full concordance with the diagnostic classifier.

### Automated adjudication enables systematic benchmarking

Automated pathogen detection enabled systematic investigations of how upstream computational parameters translate into diagnostic performance, using regression and perturbation analyses that are impractical to scale under manual expert adjudication (**Figure 6A-B**). Both contamination control and the composition of the taxonomic reference database materially affected diagnostic performance. Disabling computational contamination control caused specificity to collapse (consensus 95.4% to 88.4% without clinical notes, **Figure 6A**) while database host-genome handling traded sensitivity against specificity, with only the masked configuration used in this study achieving balanced high performance (**Figure 6B**, component-level analyses in **Supplementary Analysis**). Clinical context largely restored specificity lost to ablated filtering in the contamination control experiment but did not recover lost sensitivity in the host database experiment. This is consistent with clinical information serving as negative evidence to reject spurious detections rather than recovering pathogen signal. Regression testing of a candidate classifier update (a development build of Metabuli with syncmer support, **Methods**) showed no significant change in consensus diagnostic performance (Wilcoxon ranked-sum test, sensitive: p = 0.742, specificity: p = 0.867; **Figure S8**, **Supplementary Analysis**) demonstrating that automated adjudication makes evaluation of pipeline modifications tractable for quality assurance testing.

**Figure 6.**
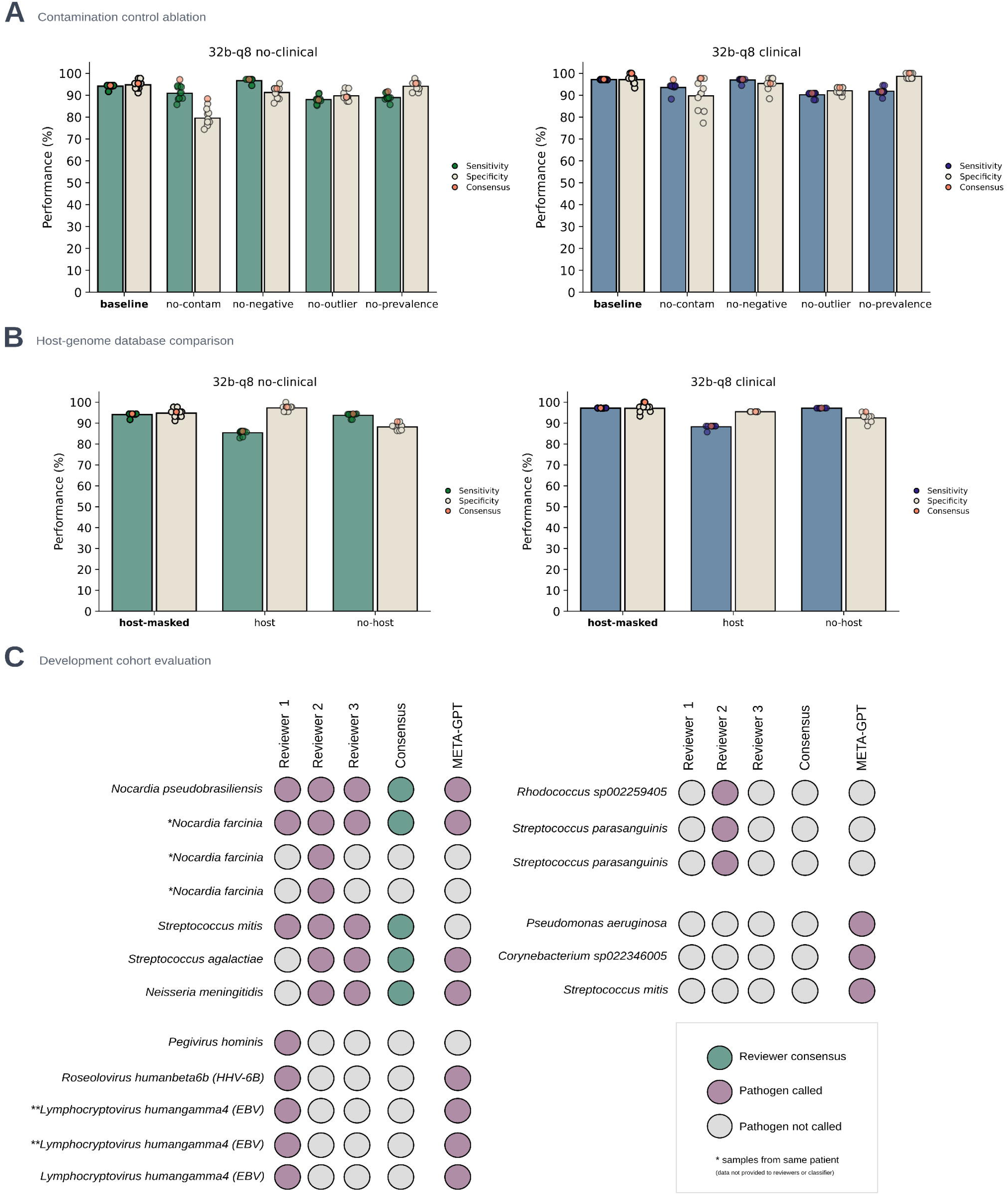
Application of diagnostic classifiers to systematic benchmarking and a retrospective development cohort. (A-B) Diagnostic performance in perturbation analyses of computational parameters for contamination control and taxonomic database composition in regards to host-genome handling for the best-performing model (qwen3-32b-q8-tiered) with (blue) and without (green) clinical notes. (C) Concordance of positive classifier and reviewer calls (red) against reviewer majority consensus (green) in a retrospective development cohort (n = 78 patients; n = 87 samples). RPM, reads per million; gray, species not called; * and ** denote detections across different samples from the same patient, information that was not available to reviewers or model during evaluation.

### Diagnostic classifier interpretation extends expert review across domains

We applied the best-performing classifier (q*wen3-32b-q8-tiered-clinical*) to a retrospective development cohort (n = 78 patients) assembled during establishment of the assay. Samples comprised suspected CNS and ocular infections without a diagnosis on routine testing, sequenced under variable, pre-production laboratory protocols and evolving contamination controls (n = 87; 70 CSF, 14 ocular fluids, 3 ocular tissues; **Methods**). Given the developmental nature of this cohort we present it descriptively rather than as a quantitative accuracy evaluation, comparing the classifier’s pathogen calls with the consensus interpretation of three clinical microbiologists (**Figure 6C, Table S6**). Reviewers each exceeded 90% sensitivity and specificity in the validation dataset evaluation, and assessed libraries individually, de-identified, and out of sequencing-run context (the same information available to the classifier) so that human and model interpretation were assessed on matched evidence. Consequently, reviewers and classifiers could not aggregate signal across multiple libraries from the same patient, including three *Nocardia farcinica* positive libraries and two Epstein-Barr virus positive libraries, which each originated from a single patient (**Figure 6C**).

Classifier calls included clinically significant pathogens not identified by routine testing (*Neisseria meningitidis, Streptococcus agalactiae, Nocardia farcinica, Nocardia pseudobrasiliensis*). The concordance matrix illustrates the value of consensus over individual interpretation, with divergences between the classifier and individual reviewers reflecting detection thresholds and prior diagnostic experience (**Figure 6C**). Classifier calls encompassed nearly every organism deemed reportable by consensus, including two bacterial pathogens (*N. meningitidis, S. agalactiae*) that one reviewer did not report, but were identified by the other two reviewers and the classifier. Conversely, the classifier missed a consensus-reportable *Streptococcus mitis* detection in one sample.

Consistent with the sensitivity-tuned filtering configuration, the classifier and individual reviewers each additionally flagged organisms that did not reach consensus. The reviewer specialising in CNS metagenomics, who did not report the two bacterial pathogens, instead flagged viral detections plausibly associated with (rare) CNS infections, including *Roseolovirus humanbeta6b* (HHV-6B), *Lymphocryptovirus humangamma4* (EBV) and *Pegivirus hominis,* recently associated with encephalomyelitis and independent CNS replication in immunosuppressed patients (Scheibe 2025). The classifier called this as a background organism of unproven pathogenicity, underscoring that emerging evidence of pathogenicity was captured by domain expertise but not by the model. Similarly, the reviewer with bacterial expertise flagged *Rhodococcus sp002259405*, noting the potential role of *Rhodococcus* species in (rare) ocular infections. Singular calls by the classifier, such as *Pseudomonas aeruginosa* and the reviewer-unsupported *S. mitis* detection arose in samples with high background contamination and low pathogen abundance (**Table S8**). *P. aeruginosa* was recovered only as a marginal call of low reproducibility (**Table S7**).

## Discussion

Metagenomic sequencing enables broad-spectrum pathogen detection for clinical diagnostics, yet its widespread adoption has been constrained by the interpretive burden associated with expert adjudication (Blauwkamp2019, Fourgeaud2024, Benoit2024, Alcolea-Medina 2025). In this study, we present a structured interpretive framework that formalizes this adjudicative process by augmenting quantitative abundance thresholds with context-aware reasoning, grounded in clinical plausibility. The framework integrates stratified taxonomic detections at species rank with diagnostic decision trees and open-weight LLMs (META-GPT), emulating the reasoning clinicians use when weighing organism abundance, contamination risk, and patient context. By directly quantifying expert adjudication performance for the first time (Chiu2019, Benoit2024, Torres-Montaguth 2025), we establish a clinically grounded benchmark and demonstrate that automated diagnostic classifiers can achieve sensitivity and specificity comparable to individual experts and consensus adjudications.

Expert review remained a highly reliable proxy for clinical truth in this study, but only in aggregate. Clinical microbiologists interpreting the same stratified evidence as the classifiers (without task-specific training or clinical notes) varied widely in individual performance. The highest-performing reviewers in the baseline evaluation all had recent hands-on experience with pathogen genomics or metagenomic data, suggesting this variability reflects familiarity with data interpretation rather than diagnostic ability. Consensus across these independent reviews nonetheless recovered near-ceiling performance. Reliable interpretation therefore depended on access to an experienced panel, or on aggregating multiple independent reviews, which may be operationally impractical for routine use (Kaur 2025). We did not formally assess the effect of training, but our proof-of-concept, model-assisted adjudication, in which experts began from a classifier-anchored starting point, markedly reduced inter-reviewer variability and raised individual performance to the consensus ceiling. However, this descriptive comparison warrants caution as reviewers were anchored on classifier calls and performed a structurally different verification task, with clinical notes made available to both reviewers and classifiers. Overall, our adjudication evaluation indicates that greater familiarity with interpreting these data would be expected to improve individual accuracy and reduce variability.

It is worth noting that the reduction in inter-reviewer variability under model-assisted adjudication is itself a manifestation of anchoring, the same cognitive bias we identify as a liability of unassisted expert review (**Appendix**). Reviewers converged in their performance because they were anchored on a common (in this case correct) starting point provided by the classifier’s pre-populated calls. This benefit relies on high anchor accuracy, a condition that was confirmed retrospectively against a reference standard in this study but cannot be guaranteed at the point of care in prospective use, where the same anchoring could reinforce classifier error. Notably, the incorrect overrides came from two reviewers who had performed strongly at baseline, each overriding a correct classifier call into an error, a manifestation of automation bias in which expert confidence displaces an accurate automated anchor rather than deferring to it uncritically. A promising direction for prospective use is therefore a human-in-the-loop design in which the classifier surfaces multiple ranked candidates rather than a single candidate, retaining the benefits of assistance while mitigating the anchoring risk identified here, an approach that warrants evaluation in prospective clinical trials.

Diagnostic classifiers enabled scalable computational benchmarking including ablation testing of contamination-control strategies, host-database configurations, and regression testing of taxonomic classification software. Our proof-of-concept analyses illustrate how technical choices translate into diagnostic outcomes. Removing in silico contamination controls substantially reduced diagnostic specificity, but inclusion of clinical notes largely restored classifier performance. Clinical context proved critical for resolving detections involving commensal bacterial organisms, enabling disambiguation that taxonomic classification alone could not achieve. These results provide computational evidence that contextualization, rather than sequence abundance alone, can determine whether an organism warrants clinical concern (Benoit 2024). We further examined how structuring taxonomic evidence either through a tiered-node (above- and sub-threshold) or a single-node (flattened representation) affected diagnostic performance and observed significant reductions in sensitivity across models, which was not restored by the inclusion of clinical notes.

Divergent effects of contamination filtering, clinical context and input structure on classifier performance are consistent with prior observations from clinical metagenomics and language-model research. Removal of contamination controls reduced diagnostic specificity by increasing exposure to low-abundance commensal and reagent-associated taxa, a dominant source of false-positive signal in low-biomass metagenomic assays (Miller2019, Benoit2024, Alcolea-Medina 2025). Incorporation of clinical notes substantially mitigated this effect, supporting the interpretation that contextual information primarily acts to suppress implausible organism calls through clinical reasoning, rather than by enhancing pathogen signal. In contrast, the marked reduction in sensitivity observed under a single-node interpretation architecture suggests a distinct failure mode driven by increased distractor burden. Prior work has shown that LLM are sensitive to both irrelevant context and input length, with excess or poorly structured information impairing reasoning accuracy and promoting conservative or unstable decision-making (Wei2022, Shi2023, Liu 2024). Stratifying detections into above- and sub-threshold tiers appears to function as an attention-shaping mechanism, constraining the hypothesis space and enabling integration of weak but internally consistent evidence. Conversely, flattening of the candidate set increases competition among marginal detections and raises the effective decision threshold for positive classification. Notably, clinical notes did not rescue sensitivity in the single-node architecture, consistent with their primary role as negative evidence for rejecting spurious detections. This suggests that explicit organization of metagenomic evidence may be critical for maintaining diagnostic sensitivity in low-biomass, noise-dominated samples.

Several important limitations of this study must be considered. First, both the validation and development cohorts were modest in size and did not capture the full diversity of clinical presentations, underscoring the need for rigorously designed prospective studies to establish generalizability and robustness. Second, the short-read sequencing protocols, computational analyses and diagnostic decision flows used here were optimized for low-biomass sample types, where pathogens must be distinguished from sparse signal amid background contamination. High-biomass samples will require alternative strategies to address signal saturation and dense microbial communities such as long-read sequencing and functional characterization (Gu2020, Alcolea-Medina 2025). Prevalence-based contamination filtering was applied on a per-run basis to account for temporally variable contamination sources, motivating future incorporation of curated contaminant databases (Miller2019, Benoit 2024).

Finally, operational and resource considerations remain substantial. Clean workflows within shared laboratory environments are essential. The use of synthetic clinical notes for spike-in controls, while necessary to evaluate contextual reasoning in non-clinical samples, may not fully capture the biases present in real-world clinical evaluations. Assessments of our model was performed on sequencing data generated from a single laboratory which may have its own unique contaminome derived from the workspace and reagents used. Therefore, further evaluation assessing performance of META-GPT on sequencing data derived from multiple metagenomic laboratories is required to derive generalisability. Furthermore, probabilistic sampling at non-zero temperatures necessitated replicate-level consensus to mitigate single-model variability. This requirement introduces nontrivial computational overhead, even with batching strategies for multi-GPU systems as implemented in META-GPT. Sustained deployment at scale will therefore require access to sufficient hardware or purpose-trained models optimized for low-memory inference. Notably, the use of locally deployed, open-weight models offers practical advantages for patient privacy and regulatory oversight by enabling full auditability of model updates and institution-level control over data governance (Sandmann2025, Tordjman 2025).

Future developments in metagenomic diagnostics and medical LLM provide clear opportunities to extend this framework from structured adjudication emulation to more robust, evidence-grounded decision support. Clinically-oriented foundation models for diagnostic reasoning could improve calibration in atypical presentations and low-prevalence settings when coupled to explicit uncertainty reporting and selective deferral to expert human review (McDuff 2025). Furthermore, retrieval- and tool-augmented language model pipelines offer a practical route to grounding interpretations in curated sources, such as pathogen references or institution-specific contaminant catalogues. As metagenomic workflows increasingly incorporate long-read sequencing and strain-resolved assemblies, downstream interpretation could move beyond “organism detected” towards clinical plausibility supported by genomic context, with different decision flows for low-versus high-biomass samples (Gu2020, Alcolea-Medina 2025). Integration of metagenomic outputs with other diagnostic machine learning models could support differential diagnoses by discriminating infectious from non-infectious aetiologies, such as autoimmune disorders or malignant neoplasms, which can mimic infectious neuroinflammatory syndromes (Ramachandran2020, Gu2021, Ramachandran 2022). Finally, prospective multi-site evaluations that combine standardized adjudication schemas and human-in-the-loop diagnostic classifiers will be essential to establish clinically meaningful benchmarks and support regulatory validation of LLM-assisted metagenomic diagnostics at scale.

Autonomous diagnostic classifiers will not replace the clinical judgment required for complex infectious disease diagnosis. In our real-world development cohort, a single classifier provided interpretive breadth across bacterial and viral disease that would otherwise require a multidisciplinary panel, yet individual domain expertise remained necessary to recognise organisms of emerging or unusual significance. By capturing the structure of expert adjudication in a form that can be systematically evaluated, diagnostic classifiers provide a scalable and reproducible foundation for diagnostic interpretation, while preserving expert oversight for organisms of uncertain clinical significance.

## Data Availability

Host-depleted sequence reads will be available under NCBI BioProject accession SUB16321535. Data used in this study is available on FigShare (10.26188/32956268). META-GPT, Cerebro, Cipher, Scrubby and Vircov are available open-source under GNU GPLv2, GPLv3 and MIT/Apache licenses (https://github.com/vidrl).

## Acknowledgements

We would like to thank staff at NECTAR / University of Melbourne Research Cloud and Victorian Infectious Diseases Reference Laboratory (VIDRL, IT) for their support with computational resources used in this study. We would like to thank laboratories at VIDRL (VI, BMP, MRL) and WEHI (Tonkin group) for providing the reference isolates in the limit of detection experiment.

## Generative pretrained transformer statement

During the preparation of this manuscript, the authors of the original draft used a locally deployed Qwen3.6-32B-A3B (QwenAI) model to rephrase and copy-edit author-written passages for clarity and concision, and transform tabular content and formatting for the LaTeX version of this manuscript. A closed-weighted LLM model was use for editing selected passages to improve readability. Authors reviewed and edited the output as needed and take full responsibility for the content of the publication. Large language models were not used to generate research data, analyses, scientific conclusions or figures, with the exception of the background element design in the schematics of **Figure 2A** and **Figure S5**. Open-weight language models that are the subject of this study (Qwen3) are described in the Methods and were used as experimental systems.

## Conflict of interest statement

The authors have no competing interests.

## Funding statement

This work was supported by the Australian Government Medical Research Future Fund (MRFF) Genomics Health Futures Mission (GHFM) Flagships - Pathogen Genomics Grant (FSPGN000045) META-GP: DELIVERING A CLINICAL METAGENOMICS PLATFORM FOR AUSTRALIA. The funding bodies had no role in the design, analysis, interpretation, or writing of this work.

## Appendix

### Precedents for expert review in metagenomic diagnostics

Expert review panels have become a central component of metagenomic assay evaluation, particularly for assessing clinical relevance and plausibility of detected organism. Two examples illustrate this role under contrasting conditions. The first is a community benchmarking challenge that isolated the interpretive step using a single clinical case (Meyer 2022). The second is a seven-year prospective implementation of metagenomic sequencing in routine clinical service (Benoit 2024). The first shows why expert interpretation is necessary for diagnosis; the second shows what is required to deliver it reliably in routine practice.

### Clinical pathogen diagnostic challenge (CAMI II)

CAMI II is a community benchmarking exercise in which independent teams analyse a shared metagenomic dataset. Its conceptual clinical pathogen challenge provided participants with a dataset derived from the blood of a patient with viral haemorrhagic fever, accompanied by an anonymised and modified clinical case description. Participants were asked to identify all pathogens present and to nominate the single organism most likely to have caused the illness. The causal pathogen, Crimean-Congo Haemorrhagic Fever Virus (CCHFV, *Orthonairovirus haemorrhagiae*), had been detected in the sample and confirmed by PCR (Ct = 27.4), although the subclinical presentation precluded definitive proof of causality. Only three of ten submissions correctly nominated CCHFV as the causal agent. The remaining seven either did not detect it or detected it without prioritising it diagnostically, despite its confirmed presence and the absence of any alternative plausible cause. Some failures therefore occurred at the point of interpretation rather than detection. This illustrated the limited clinical utility of taxonomic profiling on its own, and the substantial variability of interpretation across teams working from the same data.

### Structured expert adjudication in routine clinical implementation

A seven-year prospective evaluation of cerebrospinal fluid (CSF) metagenomic sequencing (mNGS) at UCSF demonstrates the multi-layered review process that reliable adjudication requires in routine service. The laboratory director oversaw all test approvals and interpretations. A contaminant-organism database, curated continuously from external controls over rolling three-month intervals, automatically flagged taxa for manual review, and detections above predefined positivity thresholds could be reclassified as contamination on contextual grounds, such as presence in the contaminant database, co-detection with known commensals, or evidence of cross-sample bleed. Testing requests were vetted against predefined clinical criteria, including CSF pleocytosis, biopsy evidence, immunocompromise, or subspecialist recommendation. Final diagnostic categorisation relied on retrospective adjudication by three infectious disease physicians, who independently reviewed the full clinical and microbiological workup, including mNGS and conventional results, and resolved discrepancies by consensus. Organisms were classified by clinical relevance rather than presence alone, as true positives, false positives, likely or possible contaminants, incidental findings, or findings of uncertain significance, and only those adjudicated as causative of the CNS syndrome were counted as true positives in performance metrics. Selected cases were discussed in real time with treating physicians through a clinical microbial sequencing board. This process achieved rigorous, context-aware interpretation, but it depended on sustained specialist time, curated infrastructure, and standing multidisciplinary panels that few laboratories can reproduce.

### Limitations of expert review for metagenomic diagnostics

Expert adjudication is currently the most context-aware reference standard available for evaluating metagenomic diagnostics, but it carries several limitations that constrain its reproducibility, scalability, use in optimisation, and integration into routine clinical practice.

#### 1. Resource-intensive and dependent on specialist expertise

Expert review requires substantial time commitment and depends on clinicians with expertise in both infectious diseases and the interpretation of metagenomic data. The process can involve several rounds of deliberation that incorporate clinical presentation, host factors, sample type, and sequencing results (Benoit 2024). It is therefore difficult to implement outside large, well-resourced centres with access to multidisciplinary teams experienced in metagenomic interpretation.

#### 2. Subjectivity and cognitive bias

Diagnostic interpretation is susceptible to cognitive biases, including anchoring, confirmation bias, and memory effects. When reviewers assess the same datasets repeatedly, or recall prior classifications, consistency and objectivity are reduced. These effects limit the reproducibility of expert review and reduce its suitability as a stable reference for iterative model training or optimisation.

#### 3. Incompatibility with high-throughput optimisation

Because panel adjudication is slow, it cannot be applied across the large parameter spaces explored during pipeline development. This prevents exhaustive tuning of classifier parameters, thresholding criteria, and database versions, and it precludes the use of expert adjudication as an optimisation objective in automated workflows. At present, large-scale optimisation benchmarks for metagenomic pathogen detection rely on taxonomic profiling metrics (McLaren2019, Ye2019, Meyer 2022) which do not reflect diagnostic performance in clinical applications.

#### 4. Scalability barriers in routine diagnostics

Many laboratories, regardless of size, lack the infrastructure to operate a metagenomic assay (clean space and PCR setup) and a structured expert review process. Where clinical specialists or formal review boards are not available, interpretation of metagenomic results can vary widely between operators and sites. This limits the scalability of expert-driven frameworks and introduces inconsistency in real-world use, particularly in resource-limited settings.

#### 5. Inter-reviewer variability and lack of standardisation

Even within expert panels, the criteria used to classify taxa as contaminants, incidental findings, or causative agents depend on individual reviewer experience and on the local laboratory environment, including sample-processing conditions in hospital and research settings (especially pre-analytical contamination at sample collection and concurrent enrichment-based assays that may easily contaminate shared laboratory spaces, as noted during development of the assay presented in this study). Consensus adjudication reduces disagreement, but residual variability introduces noise into performance estimates and complicates comparison across pipeline configurations and reference cohorts.

#### 6. Quality assurance and pipeline maintenance

Accredited metagenomic pipelines require formal quality assurance when software, reference databases, or algorithms change. Re-validating performance by expert adjudication after every update is impractical, and becomes more so as testing volume grows. The absence of standardised and automatable surrogate metrics complicates regression testing and makes compliance with clinical accreditation standards difficult to maintain (although certainly not impossible, as some laboratories have demonstrated).

#### 7. Retrospective benchmarking and outcome bias

Adjudicators may have knowledge of patient outcomes, which can influence classification toward concordant results. Retrospective review is also limited by missing contextual information and by the unavailability of the original reviewers. Both reduce the reliability of expert adjudication as a stable benchmark for evaluating diagnostic performance over time.

Expert clinical review remains essential for evaluating metagenomic diagnostic performance. However, the limitations described above are significant barriers to systematic improvement and wider adoption of metagenomic diagnostic assays. They are most acute where metagenomic data must be interpreted across diverse sample types, patient presentations, local pathogen epidemiology, or sequencing protocols. The present study addresses these limitations by developing a structured and interpretable framework that emulates expert reasoning using generative pretrained transformers for contextual infectious disease diagnosis and pathogen candidate selection.

## Methods

### Recruitment and sample collection

Patients and samples were recruited and collected under the META-GP research program. The META-GP program is a Medical Research Futures Fund (MRFF) funded program, established to develop a clinical metagenomic assay for Australia. Ethics approval was obtained from 8 hospitals in Victoria, Australia (HREC 74254). Known positive and known negative samples underwent passive recruitment, and residual CSF and vitreous fluid was stored for metagenomic sequencing after standard microbiological testing was performed. Most of these cases were recruited from public health laboratories. Clinical cases of meningoencephalitis or ocular inflammation, where clinicians specifically requested metagenomic sequencing to aid in the diagnosis, were actively recruited, where patient or next of kin consent was obtained. A subset of these patients was consented and recruited even with known positive or negative infections to allow access to clinical records, which was not available with passive recruitment. Samples were collected and stored at recruiting hospitals at -80°C as soon as possible. The time from collection to freezing was not documented and was not known in most cases. Samples were transferred on dry ice and then stored again at -80°C to reduce freeze-thaw cycles.

### Extraction and library preparation

200ul of CSF or vitreous fluid initially underwent sonication. Samples in 2 mL screw-cap tubes were held in a foam float so that they contacted neither the walls nor the base of the bath and sonicated in a VEVOR Professional Ultrasonic Cleaner at 40 kHz for three rounds of 5 min sonication followed by 5 min on ice. The bath was filled with room-temperature water for the validation plate; subsequent runs used ice-chilled water. Samples were then extracted using the TANBead Nucleic Acid Extractor Maelstrom 9610 (TANBead Nucleic Acid Extraction Kit OptiPure Viral Auto Tube, Cat No. W665566/W665A46) or the Qiagen EZ1 Advanced XL platform (QIAGEN EZ1C2 virus mini kit v2.0, Cat No. 955134). If 200ul of sample volume was not available, then DNA/RNA Shield (Zymo Research Cat No. R1100) was used to bring the sample up to the extraction volume. 60ul of total nucleic acid was produced half of which was combined with 70 µl of DNase master mix (RNase-Free DNase Set, Qiagen Cat No. 79256) and incubated at room temperature for no longer than 10 min. The reaction was stopped by AMPure XP bead cleanup and eluted in 32 µl of 0.1x TE buffer. This method of extraction was derived from extraction optimisation experiments (data not included). Library preparation was conducted using the NEBNext Ultra II RNA Library Prep Kit for Illumina (NEB Cat No. E7770) and NEBNext Ultra II FS DNA Library Prep Kit for Illumina (NEB Cat No. E7805), incorporating single-strand unique molecular identifiers (UMIs) (NEB Cat No. E7874 and E7416) to mitigate issues such as PCR amplification bias and barcode hopping. This approach also facilitates the accurate quantification of nucleic acids and enhances data quality for host genomic studies. Integration of UMIs required multiple steps variation to the manufactures protocol to optimise sequencing yield for low biomass samples such as CSF and vitreous fluid. In short this included the exclusion of the fragmentation step, dilution of adaptors, changes to the final PCR amplification step (19 cycles for DNA and 21 cycles for RNA) and varying the ratios of magnetic beads in the clean-up steps. Human rRNA was depleted using QIAseq FastSelect (Qiagen Cat No. 333180). Libraries were sequenced on an Illumina NextSeq 2000 using P3 reagents (300 cycles, 2 x 150 bp paired-end) with PhiX Control v3 spike-in (Illumina Cat No. FC-110-3001). The validation plate was sequenced across two P3 cartridges, each carrying a pool of 48 samples (96 libraries, comprising the paired DNA and RNA library from each sample). The plate was divided vertically by extraction platform, and pools were balanced so that each cartridge contained 24 samples extracted on the TANBead Maelstrom 9610 and 24 on the Qiagen EZ1 Advanced XL.

### Sample pooling

Library concentration was measured using the Qubit dsDNA HS Assay Kit (Invitrogen Cat No. Q32854) read on a CLARIOstar Plus plate reader (BMG Labtech) against the kit standards, and fragment size distribution was assessed on an Agilent TapeStation 4200 using High Sensitivity D1000 ScreenTape (Cat No. 5067-5584) with High Sensitivity D1000 Reagents (Cat No. 5067-5585). Standard sample pooling is performed using equimolar pooling, to derive an equal amount of reads to each sample. For CSF, sample input is not normalized due to the low biomass. Therefore, each sample gets an equal amount of reads regardless of the amount of biomass in each sample. Higher levels of host background biomass act as interference, reducing sensitivity of pathogen detection, unless sequencing depth is increased. Conversely, low levels of host background reads, sequenced at a fixed read amount can lead to over-sequencing, which in turn increases cost. We assessed whether equivolume pooling results in a dynamic method of read distribution for each sample. Illumina sequencers preferentially sequence higher concentration samples, and therefore samples with higher biomass would have a higher number of allocated reads, reducing interference and increasing sensitivity. Conversely, lower biomass samples would get fewer reads and reduce cost and over-sequencing. We spiked in Adenovirus and Enterovirus at Ct 30 and 27 respectively, into a ladder of increasing background human biomass to represent the distribution of CSF host biomass in a standard meningoencephalitis clinical cohort. The adenovirus isolate was human adenovirus 5 (ATCC VR-5, strain Adenoid 75) and the enterovirus was echovirus (*Enterovirus betacoxsackie*; ATCC VR-1867, strain Cornelis), both obtained from VIDRL and propagated on cell culture. Human background nucleic acid was prepared from Detroit 562 cells (Cat No. ATCC CCL-138, RRID:CVCL_1171; lot 70037075, supplied at passage 46), a human pharyngeal carcinoma line. The supplier certified the lot as free of mycoplasma by Hoechst DNA stain, agar culture and PCR-based assay, sterile on aerobic and anaerobic culture, negative for HIV, hepatitis B virus, HPV, EBV and CMV by PCR, and authenticated as human with an STR profile matching the reference profile for this line. Cells were pelleted, the culture medium removed, and the pellet resuspended in 1 ml of DNA/RNA Shield (Zymo Research Cat No. R1100) and vortexed. Nucleic acid was extracted on the Qiagen EZ1 Advanced XL using the EZ1&2 Virus Mini Kit (Qiagen Cat No. 955134), with 200 µl input and 60 µl elution, as for clinical samples. The background biomass DNA ranged from 1ng to 15ng, whereas RNA ranged from 5pg to 500pg. These same samples were sequenced using equimolar sequencing vs equivolume and in triplicate.

### Internal phage and synthetic controls

ERCC (External RNA Controls Consortium) RNA Spike-in mix (Thermo Fisher Cat No. 4456740) were added to RNA libraries as internal library controls. Complimentary DNA (cDNA) library internal controls (cDNA-LIC) were created for DNA libraries by reverse transcribing ERCC using the NEBNext Ultra II RNA reverse transcription steps and not performing any fragmentation during this process. Both ERCC and cDNA-LIC consist of 92 transcripts of varying length and concentrations. As part of the library preparation process, 25pg of ERCC and 25pg of cDNA-LIC were spiked into sample nucleic acid and all controls.

T4 and MS2 bacteriophages were used as internal extraction controls. Each phage lysate was cultured with specific strains of *Escherichia coli* that are susceptible to the respective phages. The mixture was then plated onto lysogeny broth (LB) agar plates using the soft agar overlay method and incubated for 18 to 24 hours at 37°C to encourage phage growth and plaque formation. If initial plating did not result in plaque formation, further amplification was attempted using the original lysate in broth culture, followed by replating. Lysates showing viable phage plaques were further cultivated. Once high-titer phage lysates (>109 PFU/mL) were obtained, the lysates underwent a cleaning process to remove any residual host nucleic acids. This was achieved by treating the lysates with DNase and RNase A to degrade any contaminating DNA and RNA, followed by a heat inactivation step to stop the enzymatic reactions. To identify the optimal concentration for spike-ins for phage internal extraction controls, a dilution series was performed with identification of the optimal concentrations of T4 phage at 3×10^8^ and MS2 phage at 2×10^6^. Internal phage controls were spiked into CSF, vitreous fluid and positive external controls prior to extraction. Both ERCC/cDNA-LIC and phage were used to assess interference of host reads and appropriateness of sequencing depth.

### External positive and negative controls

External positive controls were included alongside each sequencing run to ensure the reliability of our results. Organisms with no clinical correlation to CNS infections were chosen as positive controls, therefore, removing potential cross-contamination leading to spurious results in clinical samples. These controls consisted of the ZymoBIOMICS Spike-in Control II (Low Microbial Load Cat No. D6321), *Betacoronavirus muris* (Zeptometrix Cat No. NATMHV-ST), *Orthopoxvirus vaccinia* (Zeptometrix Cat No. 0810310CFHI), and *Saccharomyces cerevisiae* with a recombinant *Pneumocystis jirovecii* gene (Zeptometrix Cat No. NATPJI-ERC). The ZymoBIOMICS spike in Control II consists of a mix containing three bacterial strains, *Truepera radiovictrix*, *Imtechella halotolerans*, and *Allobacillus halotolerans*. All four controls were spiked into one tube containing 0.1x TE buffer. Synthetic CSF was not used due to expense, and adequate internal controls to assess for matrix effect. Exact concentrations were not known for all controls. The optimal volume for each control was determined over several sequencing experiments and attempting to normalize for genome size. The positive external controls were extracted and sequenced following the same library preparation protocols used for clinical samples.

### Limit of detection experiment

To establish the limit of detection (LOD), a dilution series experiment on a diverse panel of pathogens was performed. The panel comprised two bacteria (*Streptococcus pneumoniae* (ATCC 49619), *Haemophilus influenzae* (ATCC 49766), a fungus (*Cryptococcus neoformans*), a parasite (*Toxoplasma gondii*), a DNA virus (*Simplexvirus humanalpha1*) and an RNA virus (*Enterovirus betacoxsackie*). Viral and parasite isolates were obtained from VIDRL and the Walter and Eliza Hall Institute (WEHI): *S. humanalpha1* (HSV-1; ATCC VR-1778, strain ATCC-2011-1) propagated on A549 cells, *E. betacoxsackie* (echovirus; ATCC VR-1867, strain Cornelis) propagated on Vero cells, and *T. gondii* (type II, strain ME49) propagated on human foreskin fibroblasts. Isolate concentrations were quantified in-house by quantitative PCR on a QuantStudio 5 instrument (Applied Biosystems) and by digital PCR on a QIAcuity. Primer sequences and cycling conditions were provided by VIDRL, and oligonucleotides were synthesised by Bioneer. Each pathogen underwent a 10-fold dilution from Ct 25 to Ct 37. To mimic inflammatory CSF, mock human background nucleic acid was spiked into each extracted dilution using Detroit 562 cells. Extracted Nucleic acid was spiked at a fixed mass of 3ng for DNA and 25pg for RNA. The host DNA biomass was derived from analysis of all previously sequenced CSF samples, and was the median biomass for extracted CSF recorded from Qubit readings (data not shown). RNA host biomass is typically too low to undergo Qubit analysis and so 25 pg was chosen to represent the median biomass. TE buffer (0.1x) was chosen as the background matrix. The experiment involved library triplicate testing of DNA and RNA samples; three replicates were removed from the analysis due to failing sequencing libraries (2 x *Enterovirus betacoxsackie,* 1 x *Streptococcus pneumoniae*). Bioinformatic analysis was performed using the default Cerebro (v0.12.0) pipeline configuration and database (cipher-v2-host-masked). Nominal probit analysis was conducted across dilution-levels (Ct 25 - Ct 37) and replicates for each pathogen and classifier combination (**Figure 1**). A terminal dilution-level of Ct 38 (N/A, absolute LOD in this study) was used in all cases where reads were identified at the final dilution level (Ct 37). One set of replicates indicated operator error due to higher-than-expected sequence abundance for *Cryptococcus neoformans* (Ct 34, **Figure 1**) but was retained for analysis, as all replicates were detected in the following dilution (Ct 37). Dilutions used for library preparation were quantified by qPCR, allowing LOD₉₅ values to be expressed as approximate concentrations (copies/mL) at the nearest measured dilution.

### Analytical performance dataset

For diagnostic performance metrics, a 96-well plate was created using known-positive and known-negative clinical samples (CSF and ocular fluids), external positives, and negative controls (**Figure S3, Table S1**); CSF negatives were confirmed as autoimmune or non-infectious diagnoses based on clinical review of all CSF cases (PR, AD). Any ocular samples were considered negative if all clinical orthogonal testing was negative. Of the 87 samples, 43 were negative clinical samples, including 3 pooled negative background samples, and 34 were positive clinical samples with orthogonal testing verification, and 10 spiked samples, described below. Of these, 65 were CSF samples and 22 were ocular fluid samples. Nine control samples were included, consisting of three external positive controls, four negative template controls, and two environmental controls.

To ensure an adequate diversity of potential CNS pathogens, mock clinical samples were created for 10 cases with cultured pathogens spiked into pooled negative CSF samples. These included *Mycobacterium tuberculosis* (n = 1), *Listeria monocytogenes* (n = 1), *Aspergillus niger* (n = 3), *Orthoflavivirus murrayense* (Murray Valley Encephalitis Virus) (n = 1), and *Mastadenovirus caesari* (Human Adenovirus 5) - two samples, processed with two additional technical replicates (n = 4) (**Figure S3, Table S1**).

Discrepancy testing was performed on any sample where results of the orthogonal testing did not match the metagenomic results and where sufficient sample or extracted nucleic acid was available. This was performed in a NATA accredited laboratory on residual sample fluid, and where none was available, the extracted nucleic acid. If a sample was initially considered a known positive and metagenomics and discrepancy testing was negative for the pathogen, the sample was deemed a negative (n = 4, ocular fluids: confirmed negative for recorded *Candida albicans, Treponema pallidum* and *Staphylococcus aureus;* CSF: confirmed negative for recorded *Roseolovirus humanbeta6b* in sample with confirmed positive *Simplexvirus humanalpha2*) (**Figure S4, Table S1**). For known negative samples that were deemed positive on metagenomics, if discrepancy testing was positive, the sample was deemed a positive (n = 4, ocular fluids: *Haemophilus influenzae* in three samples, one confirmed negative in discrepancy testing for *Staphylococcus aureus*; CSF: *Streptococcus agalactiae* in one sample), and if the discrepancy testing was negative, it was deemed a negative (n = 2, ocular fluid: *Lactococcus spp.* negative in one sample; CSF: *Toxoplasma gondii* negative in one sample) (**Figure S4, Table S1**).

Due to a traceable contamination event with unknown DNA (suspected *Aspergillus flavus*) in a 0.1x TE buffer aliquot used for the external positive controls (n = 3) and one of the spike-in samples (DW-63-V55, *Orthoflavivirus murrayense*) we excluded several highly unusual taxonomic detections (*Aspergillus flavus*, *Sarcocystis neurona*, *Neospora caninum*, *Trypanosoma cruzi*, *Simplexvirus pteropodialpha2*, *Acholeplasma laidlawii*) that clustered at high abundance among these samples in the host-taxon regressions (**Figure S9**). Incidental detections were likely a result of reference genome contamination with homologous sequences from the unknown contaminant. One RNA library with a matching positive DNA virus result (*Simplexvirus humanalpha2*) failed quality control thresholds (see below) on throughput allocation and was excluded from analysis (DW-63-V03, **Figure S**).

### Reference databases and taxonomies

We developed Cipher v0.5.0 (https://github.com/esteinig/cipher) a Rust library and command-line tool for modular construction of diagnostic reference databases, allowing us to control taxonomic composition (e.g. including a human reference genome) and genome processing (e.g. masking genomes with synthetic human reads) to generate testable hypotheses regarding the impact of reference database composition on diagnostic performance (**Figure 6B**). Cipher also provides functions for creating synthetic reference panels from simulations of simplified host-pathogen metagenomes with Illumina PE (Holtgrewe 2010) and ONT signal data (Gamaarachchi 2024) (including basecalled read quality control) (Steinig 2022), allowing integration testing of reference databases to investigate potential errors arising from database composition or pipeline configurations. The database used in this study was composed from a set of reference-standard genome collections (cipher-v2, n = 130248, assemblies retrieved 31/10/2024) which included all viral reference genomes from the International Committee for the Taxonomy of Viruses (Siddell 2023) (ICTV VMR v39-2, n = 16240), archaeal and bacterial species-representative genomes from the Genome Taxonomy Database (Parks 2022) (GTDB r220, n = 113082), eukaryotic pathogen genomes from the Eukaryotic Pathogen Database (Howe 2017) (VEuPath v89, n = 657), and parasitic nematodes and platyhelminths from WormBase ParaSite (WBPS v19, n = 270). Variants of this reference collection were constructed by including the complete human reference genome (Nurk 2022) (CHM13v2, cipher-v2-host) and by masking eukaryotic and viral reference genomes using synthetic human reads derived from the human reference genome (cipher-v2-host-masked) as well as excluding contigs with masked percentage > 80%. Bacterial genomes were not masked as the process exceeded our local computational resources and the representative species genomes from the GTDB are quality-controlled a priori (Parks 2022). Cipher generated indices for aligners and classifiers used in the diagnostic pipeline with default parameters for the respective versions used in this study. The development version of Metabuli (Jaebom 2024) used in the regression testing demonstration was built with default parameters for syncmers (--syncmer). Cipher constructs a grafted taxonomy using taxonkit v0.17.0 (Shen 2021) that integrates domain-specific lineages from the taxonomic reference standards for Viruses (ICTV), Archaea and Bacteria (GTDB), and Eukaryota (NCBI Taxonomy).

### Data processing and quality control

Sequencing outputs were processed using the Cerebro v0.12.0 platform for metagenomic diagnostics in clinical production environments (https://github.com/esteinig/cerebro). Paired-end short reads (DNA and RNA libraries) with UMIs from Illumina platforms were processed in the quality control module of the pathogen identification pipeline, which: (1) aligns, estimates coverage and removes internal controls (synthetic controls for sequencing depth and biomass estimates, DNA and RNA bacteriophage for extraction verification) using minimap2 v2.28 (Li 2018) and samtools v1.21 (Li 2009) with the short-read configuration in Scrubby v1.0.0 (https://github.com/esteinig/scrubby); (2) deduplicates reads based on identical matches of the first 100 bp of a read including UMI indices if available in the sequencing protocol, optimized to reduce non-deterministic outcomes from hash collisions with fastp (Chen 2018); (3) filters reads using common quality thresholds with fastp v0.23.4 (> 50 bp, low-complexity score > 30, trimming adapters using user-provided adapter sequences, trimming poly-G reads tails > 10, trimming read tails with a sliding window and average quality scores less than Q20); (4) removes host reads by aligning against the complete human reference genome CHM13v2 using Bowtie2 v2.5.4 (Langmead 2012).

For each library, we derived a run-level quality summary and classified assays into three categories (*fail*, *pass-ok*, *pass-good*) using pre-defined numeric thresholds on read depth, control performance and base quality empirically determined during the development phase of the assay (validation plate quality control matrices: **Figure S9-S10, Figure S13-S15**). Libraries with < 1×10^6^ input reads, < 1 x 10^4^ non-host (output) reads, < 40 detected ERCC constructs or phage control genome coverage < 40% (T4 DNA or MS2 RNA) were classified as *fail*; libraries with 1 ×10^6^ - 5×10^6^ input reads, 1×10^4^-1×10^5^ non-host reads, 40-60 detected ERCC constructs or phage coverage 40-60% were labelled *pass-ok*; and runs exceeding 5×10^6^ input reads, > 1×10^5^ non-host reads, > 60 ERCC constructs and phage coverage > 60% were classified as *pass-good*. Base quality metrics from fastp were evaluated analogously, with libraries failing if < 80% and < 70% of bases were Q20 and Q30, respectively, considered *pass-ok* in the ranges 80-90% and 70-85%, and *pass-good* if ≥ 90% and ≥85% of bases were above Q20 and Q30, respectively; excluding control libraries, aggregate quality status was set to *fail* if more than one metric failed, *pass-good* if all but one metric was *pass-good*, and *pass-ok* otherwise. In practice, a tiered quality scheme separates runs that are genuinely non-interpretable from those that can detect true positives but lack the power to confidently exclude infection, which is essential in low-biomass diagnostics where false negatives carry substantial clinical risk. *Pass-good* libraries permit high-confidence negative calls, *pass-ok* libraries allow actionable positives but only qualified negatives, and *fail* libraries are withheld from interpretation or scheduled for repeat sequencing.

### Pathogen detection and panviral enrichment

Multiple strategies for read classification and profiling against the Cipher database were implemented for pathogen detection at species rank: (1) K-mer classification and profiling in default configuration uses (a) Kraken2 (Wood 2019) with default parameters followed by Bracken (Lu 2017) corrections at species rank with a minimum of 3 classified reads and 150 bp read length configuration, (b) Metabuli v1.0.5 (Jaebom 2024) with default parameters in non-precision mode for short reads, and Metabuli v1.1.0-dev with syncmer support, (c) ganon v2.0.1 (Piro 2025) in binning mode (sequence abundance) with the default expectation-maximization algorithm for multiple match resolution at species rank; (2) Metagenome assembly of short reads in default configuration uses MEGAHIT (Li 2015) with k ∈{21, 29, 39, 59, 79, 99, 119, 127} and a minimum assembled contig length of 200 bp, followed by contig alignments using nucleotide BLAST (Althscul 1990) at a minimum reporting threshold of 80% nucleotide identity, e-value threshold of 1e-06 and highest bitscore alignment for taxonomic identification among multiple matches; (3) Read alignment was implemented only for the viral database component (ICTV) due to excessive memory and runtime requirements to build complete database indices, which exceed our available computational resources. Vircov (https://github.com/esteinig/vircov) was deployed with default parameters for Bowtie2 alignment and coverage estimates using a supervised binning strategy from taxonomic annotations of reference sequences at species rank. It has previously been used for the recovery of viral genomes from rapid antigen test devices (Moso 2024). Cerebro implements standalone pipelines supporting the main protocol, including a viral enrichment workflow with configurations suitable for panviral probe-hybridisation capture panels, and a workflow for segmental aneuploidy detection (Talevich 2016) from host background DNA for differential diagnosis of malignant neoplasms that may be detectable in CSF and mimic neuro-inflammatory symptoms (Gu2021, Ramachandran 2020).

### Platform modules and data processing

Cerebro is a computational stack, comprising a server (cerebro-server) that provides the access programming interface (API) with role-based authentication and data access for a document-oriented database, a command-line client application (cerebro-client) and a team-based, multi-tenant user interface (cerebro-app) for data management and interpretation of metagenomic outputs (**S12**). Unified data models are constructed and stored for each sequenced library, which includes metadata on the pipeline, protocol configurations, tags for identification of controls (NTC, ENV, POS) and nucleic acid libraries (DNA, RNA), information about the biological sample it derives from, and unified evidence from the pathogen detection outputs. Raw data outputs from the quality control and pathogen detection modules of the core Nextflow (DiTommaso 2017) pipelines (see above) are parsed into a custom database model with standardized taxonomic lineages based on the reference taxonomy from Cipher (cerebro-pipeline). During this process, evidence from each of the alignment, k-mer classifiers and metagenome assembly results is aggregated for each taxon at each rank and stored with reference to its taxonomic lineage (d ;p ;c ;o ;f ;g ;s). As primary abundance, evidence records use measure depth normalized read counts (reads-per-million, RPM) for k-mer classifiers and viral alignments, as well as the number of bases (sum of all contig lengths) produced by metagenome assembly. In the context of our database scheme, taxon abstractions with evidence records enable database queries and aggregation pipelines to process and filter operational taxonomic units in reference to other libraries in the collection, for example to retain classifier-specific evidence in comparisons against negative template controls (see below).

### Contamination controls and filters

To mitigate environmental contamination and reagent-derived artifacts, we implemented a batch control-based filtering strategy that compares taxon-specific read support in negative controls to that in biological samples. For each taxon, Cerebro aggregates the abundance signal (RPM) across all negative template controls (NTCs) and environmental swabs (ENVs) for the batch and nucleic acid library type (DNA or RNA). Evidence entries for a taxon are retained only if the signal in the sample exceeds that in the controls by a user-defined ratio threshold. Formally, a taxon-specific evidence record is retained if:

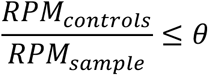

where θ is the filter threshold (default θ = 1). This ratio is evaluated independently for each classification tool and abundance mode (Sun 2021) (e.g. classification with Kraken2 vs. profiling with Bracken; in this study all methods use taxonomic read classification). By discarding evidence that is more abundant in controls than in the sample, this filter reduces the risk of false-positive detection due to systemic background noise. Negative control comparisons interact with the ensemble evidence threshold filter (see below) where evidence from specific classifiers can be removed and can lead to filtering by ensemble requirements (e.g. by requiring at least three k-mer classifiers with any RPM evidence). In the context of our ensemble approach, this evidence specific negative control parameter replaces the single-classifier threshold ratio against negative controls (RPM-ratio) in previous studies (Miller2019, Benoit 2024).

To suppress spurious taxonomic signals resulting from low-level environmental contamination (e.g. reagent, operator, laboratory, hospital, sampling site, spillover) or reference assembly contamination, we implemented a batched prevalence contamination filter. This filter evaluates all sample libraries within a batch (in this study per sequencing run) stratified by nucleic acid type (DNA or RNA) and excludes taxa that exceed a predefined prevalence threshold across the given batch of samples. Specifically, for a given taxon, we count the number of samples in which it exceeds a minimum abundance threshold (default: RPM > 0) and compute its prevalence as the proportion of libraries in which it is detected:

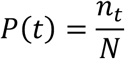

where nₜ is the number of libraries with taxon t above the RPM threshold, and N is the total number of libraries in the sample batch. Taxa with P(t) ≥ τ, where τ is the user-defined threshold, are excluded (default τ = 0.6 for taxa occurring in ≥ 60% of libraries). The rationale is that taxa appearing at low abundance in the majority of samples are likely artifacts of systemic contamination, for example from environment, reagents or reference assembly.

Notably, samples identified as outliers via regression analysis can be excluded from this prevalence calculation to avoid masking genuine taxon presence due to contamination artifacts, for example low-level host background reads retained in the sample mapping to host contamination artifacts in database reference genomes. We implemented a regression-based approach (Zinter 2019) to identify and exclude affected samples from prevalence-based contamination filters. Specifically, we model the relationship between human RPM (x) and taxon RPM (y) across all libraries in a sequencing batch using linear regression on log-transformed data:

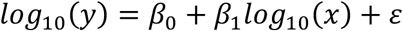

While the expected relationship is typically neutral or negative, a notably positive slope may indicate reference genome contamination with human sequences (e.g. reference assembly contamination of *Toxoplasma gondii* in the unmasked database cipherv2-host, **S13**). We define 95% prediction intervals using the Students-t distribution around the regression line and classify samples as outliers if their log-transformed taxon RPM exceeds the upper bound of this interval (**Figure 2**). Outliers, which show greater-than-expected taxon signal relative to host background are presumed to reflect genuine taxonomic presence and are therefore excluded from the prevalence filter, which would otherwise suppress taxa appearing in the majority of libraries across the batch due to remaining background reads. Host RPM are therefore utilized as a proxy for total nucleic acid content of the sample and the regression is applied to DNA libraries only (primary tier), as these are not depleted in the laboratory protocol (unlike RNA libraries; **Figure 2; Methods: Extraction**) and must have sufficient abundance (i.e. not applicable to secondary or target tiers).

### Tiered threshold reporting and configuration

We implement a tiered thresholding framework to integrate taxonomic evidence from multiple classification and profiling strategies stratified by confidence in abundance and congruence among classifier ensembles (McIntyre2017, Ye 2019). This formalizes and emulates interpretation of sub-threshold and high-priority detections that are usually at the discretion of expert adjudication (Benoit 2024). Read-level abundance (RPM) and assembly-level metrics (assembled bases) serve as primary detection metrics. Filters are applied in a staged process: a global evidence filter is applied to all taxa (see also global abundance thresholds, **Figure 2**), followed by domain-specific filters applied to eukaryotes (d Eukaryota), prokaryotes (d Bacteria, d Archaea) and viruses (d Viruses). In the first stage, application of the global filter enforces evidence inclusion criteria based on taxonomic rank (in this study: species rank) and classifier ensembles, as well as read-and assembly-level metrics. If a particular tool does not meet the criteria, its evidence entry in the ensemble data model is removed. In the second stage, lineage-aware filters require taxa to be supported by evidence from DNA or RNA libraries with a minimum number of detection tools per strategy (alignment, k-mer, assembly) above specific abundance thresholds, including distinct alignment regions at low genome coverage (Miller2019, DeVries 2021).

Stratified threshold configurations are then applied to create three reporting tiers: **(1)** *Primary* taxa (above-threshold) pass strict evidence criteria across multiple tools and modalities and represent high-abundance, high-confidence pathogen detections; **(2)** *Secondary* taxa (below-threshold) pass more lenient criteria (lower abundance thresholds, fewer tool concordance for viruses) and represent plausible but less abundant calls; **(3)** *Targeted* taxa include high-priority pathogens at low-abundance restricted to a curated list of pathogen targets, for example, viruses infecting vertebrates, syndromic pathogens of interest or cohort-specific targets. Any taxon in the primary tier is excluded from the secondary tier, and any taxon in the primary or secondary tier is excluded from the target tier (**Figure 2**). This tiered filtering system accounts for differences in genome composition and modes of infection across evolutionary distinct organisms, as well as technical bias in laboratory protocols and computational tools extensively reported in previous studies (McLaren2019, Ye2019, Meyer 2022). Expert review usually first considers above threshold taxa and then, if no plausible pathogen is detected, subthreshold taxa (below-and target-thresholds).

In the default filter configuration (**Figure 2**), we consider evidence for taxa at species rank (s) without global abundance thresholds from viral alignment (Bowtie2 + Vircov), k-mer (Kraken2, Metabuli, ganon2), and assembly strategies (MEGAHIT + BLAST). In the primary tier configuration, viral taxa are required to have evidence from either DNA or RNA libraries with alignment RPM ≥ 10 and three k-mer classifier with RPM ≥ 10; archaeal, bacterial and eukaryotic taxa are required to be detected in DNA libraries by at least three k-mer classifiers with RPM ≥ 10; assembled contigs with any number of bases are additionally required for eukaryotes. This reflects a deliberate trade-off, a stringent criterion that reduces eukaryotic sensitivity (consistent with the lower assembly-based sensitivity in the limit of detection experiment, **Figure 1**) but suppresses spurious species-level calls arising from contamination and host-homologous regions in eukaryotic reference collections, which we observed to surface frequently during assay development in the absence of an assembly requirement. In the baseline secondary tier configuration, viral taxa are required to have evidence from DNA or RNA libraries from ≥ 2 k-mer classifier with RPM ≥ 3; archaeal and bacterial taxa are required to be detected in DNA libraries by at least three k-mer classifiers with RPM ≥ 1; eukaryotic taxa are required to have evidence from DNA libraries and ≥ 3 k-mer classifiers with RPM ≥ 5, plus assembled contigs with any number of bases. In the baseline target tier configuration, only high-priority viral taxa infecting vertebrates are considered (ICTV host annotations) and require evidence from DNA or RNA libraries, ≥ 1 k-mer classifier as well as ≥ 1 alignment classifer with ≥ 2 distinct regions of alignment, when whole genome coverage is ≤ 20%.

Additionally, we provide an option to display only the species with the most supporting evidence if multiple species from a single genus (default: ≥ 5) are detected for prokaryotes (d Archaea, d Bacteria) and eukaryotes (d Eukaryota) (**S12**). This was based on observations from our data and a previous study (Gu 2020) that when high abundance species from these domains are taxonomically identified in short-read sequence data, an excess of species in the same genus are often identified at the same time. This can make the tiered interpretation display more difficult to interpret and evaluate for less abundant species. We compute a best-supported species score S(s; w) by first summing all read-based evidence (RPM) from supporting classifiers (profile records, *P*ₛ) for each species s into Rₛ:

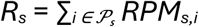

and all evidence from assembled contigs into Bₛ:

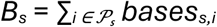

The best-supported species score S(s; w) adds these as:

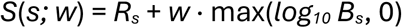

where *w* is a weight that scales bases to the RPM range (default: *w* = 1.0). Within each genus *g* that belongs to the allowed domain set G_D_ and has at least *m* species, we then select the single species s_g_* that maximizes S(s; w):

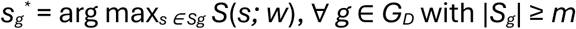

Genera outside the domain set, with too few species or taxa above genus rank are left unchanged; activation of best-supported species display filter is optional in the application interface.

### Expert adjudication evaluation

Baseline expert performance was established through the Cerebro tiered-threshold training interface (v0.12.0) which presents each sample with the same stratified taxonomic evidence (above-threshold, below-threshold, target tiers) and contextual information used in the prompts for the diagnostic classifiers. Clinical microbiologists unfamiliar with the validation dataset and given no task-specific training (n = 10) independently adjudicated each sample by selecting the single most likely pathogen from the displayed taxa, or by recording the sample as negative (non-infectious) where no plausible candidate was present. The dataset was shuffled once and presented in the same randomized order to every reviewer. These independent, unassisted adjudications estimate standalone expert performance and its between-reviewer variability and constitute the human benchmark against which the classifiers were compared (**Figure 3**).

To evaluate model-assisted adjudication, the reviewers who had completed the unassisted baseline repeated the exercise after an interval of approximately six months to attenuate recall of individual samples (n = 8 of 10 baseline reviewers). The dataset was shuffled once and presented in a common order, and reviewers now performed a correction task in which the diagnosis proposed by the diagnostic classifier (qwen3-32b-q8-tiered-clinical) was pre-populated: for samples called infectious the selected pathogen was highlighted as positive in the taxon table, and samples called non-infectious were pre-assigned negative. Clinical notes were provided to both reviewers and the classifier. Reviewers either endorsed the proposed call or overrode it by reassigning the diagnosis or selecting a different pathogen.

Because the correction task anchors reviewers on the classifier’s pre-populated call, the resulting adjudications are not independent of the model and are not directly comparable to the unassisted baseline as an estimate of standalone expert performance; the exercises also differ in task structure (de novo selection versus verification). The assisted exercise was therefore interpreted as a measure of expert endorsement of, and disagreement with, the classifier’s outputs. As the reference classifier reached near-ceiling performance on this dataset (its only missed detection, *Cryptococcus gattii* was removed by filtering and presented as negative) essentially all its visible calls were concordant with the orthogonal reference. Overrides therefore represented reviewer disagreement with calls that were already correct, and the exercise assessed whether expert oversight preserved or degraded the performance of a strong classifier (as opposed to correcting a weak classifier).

For each reviewer we recorded the endorsement rate (the proportion of samples for which the final call matched the classifier’s proposed diagnosis, and, for infectious calls, its proposed pathogen) and the override rate and its effect on accuracy relative to the reference classification. Performance of the assisted workflow was summarized as the sensitivity and specificity of the final adjudications and reported as a human-in-the-loop system. The effect of assistance on between-reviewer variability was assessed by comparing the dispersion of individual performance between the unassisted and assisted exercises (**Figure 5**). Because some reduction is expected when reviewers are anchored on a common output, this analysis emphasizes whether accuracy was maintained rather than the magnitude of variance reduction alone. This evaluation was opportunistic in scale, constrained by the limited availability of clinical microbiologists for repeated structured adjudication, and is presented as a demonstration of model-assisted adjudication rather than a formally powered reader study.

### Diagnostic classifier design and implementation

We based the initial design of our diagnostic classifiers on emulating expert review and candidate selection in the tiered threshold structure presented in Cerebro. Here, experts are encouraged (by design) to first check the above-threshold taxonomic identifications and if no candidate is found the sub-threshold sections (below-threshold and target categories). If no taxonomic identifications are retained by any filter, the sample is automatically assigned ’Non-Infectious’ (negative). We modelled this structure using configurable decision trees composed of labelled nodes that fetch taxonomic identification results and prompt the models to make a diagnosis. Each tree includes decision nodes for above-threshold or sub-threshold categories and terminal nodes for classification (’Infectious’ or ’Non-Infectious’). If the ’Infectious’ terminal node is reached, the models are prompted to select the most likely pathogen candidate from the detected organisms. For the purpose of this study, two simple topologies were implemented: a tiered-node tree that emulates expert review (above-threshold first, followed by sub-threshold) and a single-node variant that evaluates all tiers together (above-threshold together with sub-threshold). Each node generates a standardized prompt containing the following sections: [System] (system prompt), [Context] (assay information, sample type, clinical information), [Data] (threshold-specific taxa with evidence), [Tasks] (diagnostic objectives), and [Instructions] (output formats) specifying machine-readable outputs for label determination and species names for pathogen selection (‘pathogen’, with option for multiple ‘candidates’) to be parsed from the generated text. Prior to prompting, taxa may be display-filtered in order to retain the best-supported species (matching the option during expert review in the interface, default: > 5 species for prokaryotes and eukaryotes) or exclude phage from prokaryotic host annotations from ICTV (default: off). Diagnostic classifiers log node-level memory (inputs, prompts, answers, and parsed outcomes) and iterate within a bounded repeat limit (3 attempts maximum by default) when a decision cannot be extracted from the generated text (failure to recognize or generate output tags deemed ‘Indeterminate’), proceeding along *true*, *false*, or *next* edges until a final diagnostic label is reached (‘Infectious’, ‘Non-Infectious’, ‘Indeterminate’).

We implemented on-device generation with the candle stack (https://github.com/huggingface/candle) using quantized weights and a minimal abstraction over model families (Qwen series). Prompts are wrapped with model-specific control tokens, then tokenized and streamed through a two-phase loop: an initial prompt pass, followed by an autoregressive sampling loop. Sampling uses candle’s LogitsProcessor (temperature with top-K/top-P) and a configurable repeat penalty over the last n tokens. Outputs are decoded online with a token stream and, when present, thinking traces are split from the final answer. Qwen3 models were run with recommended parameters in thinking mode (temperature: 0.6, top-P: 0.95, top-K: 20) and non-thinking mode (temperature: 0.7, top-P: 0.8, top-K: 20) with a maximum output sampling length of 20000 tokens and a repeat penalty of 1.1 within a 64-token window. All model prompts were configured with the default system prompt, assay context, task and instruction sections (**Figure S4**). Peak GPU memory usage was measured per batch of samples from the validation plate (n = 12) which were run sequentially on each of the eight NVIDIA A100-SXM4-80GB accelerators in our local cluster using a polling function (100 ms) and the NVML library wrapper for Rust (https://github.com/rust-nvml/nvml-wrapper). Runtime was measured per sample, starting before and stopping after execution of the inference pipeline that produces the diagnostic results for each sample. Local generation ensured that no information used in the prompts and generated text (including clinical notes) left the institutional compute environment; no data was sent to remote providers or left the local network.

### Orthogonal testing and performance evaluation

Each diagnostic interpretation (whether produced by an individual reviewer, a single model replicate, or a consensus) was scored against the orthogonally confirmed reference result of the sample to populate a per-sample confusion matrix. A sample classified as infectious was recorded as a true positive only when the selected pathogen matched a taxon defined as positive by orthogonal testing at species rank. An infectious call naming a non-matching or unspecified organism was recorded as a false positive. A sample classified as non-infectious was scored as a true negative or false negative according to the reference status. Samples without an interpretable orthogonal result (model failure, “Indeterminate’), control libraries, and detections not retained (orthogonal reference detection < experimental LOD, where PCR testing was available) were assigned indeterminate (IND), control (CTRL), or not-considered states (NA) respectively and excluded from fixed-point performance metrics. From the retained counts, sensitivity, specificity, and positive and negative predictive values were computed using fixed point metrics at the final decision level as:

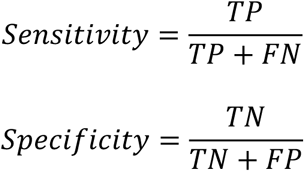

where TP, TN, FP, and FN denote the total number of true-positive, true-negative, false-positive and false-negative diagnoses across the scored set. These fixed-point metrics were used because the diagnostic pipeline does not rely on a single, unfiltered, continuous score to which a threshold can be applied, such as those generated by the taxonomic classifiers used in the pipeline; instead, it applies compound decision rules composed of multiple variables, tiered thresholds, and logical operators, including pipeline-level configurations and classifier ensemble thresholds, to arrive at a single diagnostic label. Consequently, taxonomic classification outputs are not rankable across a continuous scale, precluding threshold-dependent evaluations such as Receiver-Operating Characteristic (ROC) curves and Area Under the Curve (AUC) scores.

Consensus diagnoses were obtained by majority vote across the replicates of a model configuration or across independent expert reviewers. Assigning *n*_+(*s*)_ and *n*_–(*s*)_ for the number of assessments classifying sample *s* as infectious and non-infectious respectively, the sample was assigned an infectious consensus when *n*_+(*s*)≥*n*−(*s*)_, with ties resolved in favor of an infectious classification to prioritize sensitivity in this low-biomass setting. For consensus-positive samples, the consensus pathogen *p* (*s*) was taken as the most frequently selected species among the infectious assessments:

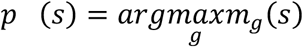

where *m*_g_(*s*) is the number of infectious assessments of sample *s* that selected species *g*. The resulting consensus interpretation was then scored against the orthogonal reference identical to individual interpretations to yield consensus performance metrics.

### Review board, clinical notes and reporting

Initital sequencing results of the validation dataset were reviewed by an expert panel consisting of at least one clinical microbiologist (KB, CKL), bioinformatician (ES), laboratory microbiologist (MK, KD) and neuro-infectious diseases clinician (PR, AD). Quality control metrics and abundance-ranked taxa with their ensemble classifier evidence were reviewed in the Cerebro application (v0.10.0) and were assessed for library quality, presence of compatible organisms, and confidence in results, including considerations of possible contamination events, sources of computational uncertainty and orthogonal testing data where available. Clinical case information was presented where relevant. For evaluation with experts and classifiers, this information was compiled by a neuro-infectious diseases physician from the records database (AD) and stripped of any references to infectious organisms (e.g. orthogonal tests for infectious organisms) to avoid injecting anchoring bias into the model prompts; clinical notes were otherwise allowed to contain references to orthogonal tests (e.g. blood or MRI results). Synthetic clinical notes were prepared by another neuro-infectious diseases clinician (PR) with no reference to existing cases (**Table S1**). Clinical reports can be created in the Cerebro application using a WebAssembly reporting library built around Typst v0.12.0 (https://github.com/typst/typst) which ensures that patient information is not transmitted from the local network where the reports are generated (**S12**). A report is generated for a single pathogen based on expert review of the taxonomic identifications at species rank. Reports were loosely based on a previous design from antimicrobial resistance profiles in *Mycobacterium tuberculosis* (Crisan 2018) and are compliant with relevant sections of ISO 15189:2022.

### Retrospective development cohort evaluation

We retrospectively analysed a dataset of CSF and ocular fluids (as well as three ocular-tissue samples, n = 118) recruited and sequenced during development of the assay when clean laboratory workflows and contamination controls were being established (2023-2024) (**Table S5**). Several samples were excluded due to quality and traceable contamination events affecting specific runs: *S. pneumoniae* contamination from high biomass run controls in the initial stages of the assay protocol (n = 11); low-level viral amplicon contamination from parallel enrichment work conducted in the laboratory at the time (n = 22); and insufficient sequencing depth (n = 5) in either DNA or RNA library, where additional sample was not available for re-sequencing. We additionally excluded any taxonomic identifications for the genus *Orthopoxvirus* due to low-level contamination from inclusion of high-abundance *Orthopoxvirus vaccinia* in the positive controls and spill-over or environmental contamination (later samples) as well as amplicon contamination from urgent outbreak response work in the laboratory during the global emergence of *Orthopoxvirus monkeypox* (earlier samples). Known bacteriophages were excluded from the filtered taxonomic identification results using ICTV host annotations (one sample, DW-63-310, *Pahexavirus lauchelly*). RNA libraries from a single run with failing RNA barcodes were repeated and analysed separately from the corresponding DNA libraries (n = 21). Laboratory protocols and library quality were therefore highly variable in this cohort, in some cases preceding the addition of unique molecular identifiers (UMI) and bacteriophage spike-ins for DNA and RNA libraries (**S14-S15**). Patients in this cohort were negative for infectious diseases on routine orthogonal testing prior to being enrolled in this study (**Methods: Recruitment**). This cohort therefore represents a scenario where a metagenomics assay is being established in a shared laboratory space while patients with unknown diagnoses were recruited for metagenomic sequencing as a broad-spectrum diagnostic test. The final dataset (n = 79 patients, n = 87 samples, where n_CSF_ = 70, n_Ocular_ = 14, n_OcularTissue_ = 3) was retrospectively analysed alongside samples and controls from their respective sequencing runs to ensure that batch contamination filters and negative controls were applied as if the data was sequenced in production. In the tiered filter, we used the sensitive default configuration but additionally specified a minimum RPM ≥ 3 for bacterial/archaeal lineages as this dataset was generally more contaminated with background taxa due to the protocol changes and contamination control establishment during development/

### Statistical tests and replicates

We applied non-parametric paired tests to assess differences in model-derived diagnostic performance across parameter settings and replicates. Each parameter setting was run ten times using identical inputs but stochastic sampling to capture inherent model variability driven by probabilistic sampling (non-zero temperature with a fixed seed, and recommended temperature settings for thinking and non-thinking modes). Replicates therefore provide within-condition variance estimates. Paired non-parametric tests were then applied across matched replicates between parameter conditions to determine whether observed performance differences exceeded the stochastic variability of the model. For continuous performance metrics (sensitivity, specificity) computed per sample block, Wilcoxon signed-rank tests for single conditions without correction for multiple testing (**Figure 4C-D, Figure S8**) implemented with scipy v1.16.2 (Virtanen 2020) for Python. In these comparisons, we evaluate whether observed performance differences arise from parameter variation (e.g. clinical notes, reasoning or software versions) rather than stochasticity in the autoregressive sampling loop (non-zero temperature).

We note that replicates of interpretation by individual clinical microbiologists on the same dataset were conceptually inadequate for statistical comparisons, as repeated interpretation of the reference dataset would not have been independent due to memory bias (which we attempted to mitigate for the assisted interpretation task by conducting it six months after the baseline interpretation). This was not the case for the diagnostic classifiers, which do not retain a memory of previous applications to the same reference dataset and can be run independently. Therefore, replicates from different clinical microbiologists were collected for comparison, reflecting realistic expert review and consensus diagnoses processes employed in clinical practice, such as by convening review boards (Benoit 2024).

We used McNemar’s test to assess paired diagnostic *consensus* outcomes between parameter configurations. McNemar’s is appropriate for evaluating differences in positive/negative result agreement on the same samples (Benoit 2024). For sufficiently large numbers of discordant pairs (≥ 25), the test statistic is approximated using a chi-squared distribution with 1 degree of freedom. However, when the number of discordant samples is small (< 25), the standard test using the chi-squared approximation becomes unreliable.

McNemar’s exact variant, which uses the binomial distribution under the null hypothesis that both classifiers have equal error rates, is commonly applied in these cases. However, the exact test is known to be overly conservative, especially for small sample sizes, often failing to detect true performance differences (Fagerland 2013). We therefore use the mid-*p* McNemar’s test, a refined version that adjusts the exact p-value to reduce conservativeness, maintaining type one error control while offering increased statistical power (Fagerland 2013). Specifically, it subtracts half the probability mass of the observed outcome from the exact p-value (Fagerland 2013). This refined approach is well suited in settings where sample sizes are limited, and discordant cases are few (which is the case for our validation dataset). However, in the context of this study, even small differences in sensitivity or specificity are clinically meaningful, as they involve critical conditions, such as suspected meningoencephalitis in our validation dataset and development cohort. McNemar’s test results were therefore integrated with manual false-negative audits and evaluation (**Figure 3C, Figure S7**).

### Data visualisation

Individual plots throughout this study were produced in Python and Rust and exported as vector graphics. Figure layout, panel arrangement (including panel letters and header text) and proportional resizing were performed in Inkscape v1.2.2. No adjustments were made that alter the data represented in any panel. Sequential numbering of reviewers (R1-R8) in Figure 5A were adjusted manually to reflect the individual reviewer identity consistent with Figure 3A (data available in **Table S1**). The tiered-filtering workflow schematic (**Figure 2**) and classifier architecture (**Figure S3**) are explanatory diagrams assembled from background elements (see generative pretrained transformer statement) and edited in Inkscape v1.2.2 to illustrate the stratification logic and node architecture described in the **Methods**. Species annotations to the right of the override matrix (**Figure 5**) were added in Inkscape v1.2.2 (data available in **Table S1**). The development cohort dot-matrix (**Figure 6**) displays results available in **Table S6** and was drawn in Inkscape v1.2.2. Validation dataset and ocular sample subset diagrams (**Figure S3, S4**) were drawn in Excalidraw v0.18.1 and subsequently modified in Inkscape v1.2.2 (data available in **Table S1**).

## Supplementary Analysis

### Impact of reasoning on diagnostic performance

Disabling explicit reasoning in the Qwen3 models (“thinking”) resulted in a consistent reduction in diagnostic sensitivity, despite equivalent inputs and inclusion of clinical notes (**Figure 4C**). This indicates that the primary contribution of reasoning in this setting is not signal discovery but structured integration of weak or borderline evidence. Prior work showed that chain-of-thought prompting improves performance on multi-step reasoning tasks by eliciting intermediate reasoning steps that support structured evidence integration, rather than altering the underlying input signal (Wei 2022). Explicit reasoning supports rejection of the null hypothesis when evidence is internally consistent by facilitating structured integration of partial cues, effectively lowering the functional threshold for positive classification and improving recall under uncertainty (Kojima2022, Wang2024, Sim 2025). Explicit reasoning pathways also improve self-verification and diagnostic commitment in medical language models (Kim 2025). In contrast, non-reasoning inference defaults to more conservative or poorly calibrated pattern completion in ambiguous clinical contexts. More broadly, these findings align with recent evaluations of clinical foundation models indicating that deliberative reasoning layers primarily enhance diagnostic commitment and robustness through improved integration of existing clinical information, rather than expanding the underlying evidence signal available to the model (McDuff 2025).

### Taxonomic database composition affects diagnostic performance

Because the host-depletion step in the quality control module of the pipeline (Bowtie2 against CHM13v2) favours specificity over sensitivity i.e. retaining residual host reads rather than risking removal of genuine pathogen reads (optimized for low-microbial biomass samples) the composition of the taxonomic database must itself account for the human reads that survive depletion. We compared three database (cipher-v2) configurations for the best performing classifier (qwen3-32b-q8-tiered): exclusion of the human genome (no-host), inclusion of the human genome (host), and inclusion with masking of viral and eukaryotic references by synthetic host reads (host-masked, used throughout this study). Relative to host-masked, excluding the human genome left sensitivity unchanged (consensus 94.4%) but reduced specificity (95.4% to 90.7% without notes) consistent with residual host reads being misclassified as taxa and causing false positive diagnoses (**Figure 5B**). Conversely, including the human genome without masking of viral and eukaryotic references reduced sensitivity for pathogen detection (94.4% to 86.1% without notes) while preserving specificity. Clinical notes did not rescue either configuration (host: consensus sensitivity 88.6% with notes below the 97.2% ceiling; no-host: consensus specificity 95.4% with notes below the 100% ceiling; **Figure 6B**) indicating that the degradation arises from taxonomic misclassification and filtering upstream of interpretation, where clinical reasoning cannot recover reads that were never attributed to the pathogen. Only the host-masked configuration achieved balanced high sensitivity and specificity, overall suggesting that host-related database composition can have a substantial effect on diagnostic performance (with the classifiers used in this study).

### Contamination-control components act on distinct performance axes

Contamination control was assessed by disabling all contamination controls in the tiered ensemble filter (no-contam) and by removing its individual components: the negative-control comparison (no-negative), the batched prevalence filter (no-prevalence) and the regression-based outlier rescue that protects genuine taxa from prevalence filtering (no-outlier) (**Figure 6, Figure S3**). Disabling all contamination control preserved sensitivity but caused specificity to collapse and become more variable across replicates (consensus 95.4% to 88.4% without notes; replicate-level specificity 94.7% to 79.5%) consistent with low-level commensal and reagent-associated taxa dominating false-positive signal in low-biomass samples. The individual components acted on distinct axes: removing the prevalence filter reduced sensitivity while leaving specificity unchanged (94.4% to 91.7%), consistent with an increased burden of contaminant taxa promoting more conservative pathogen calls. Removing the outlier rescue produced the largest single-component degradation, lowering both sensitivity and specificity (94.4% to 87.9% and 95.4% to 89.1%) as genuine taxa were over-aggressively removed by prevalence filtering. Removing the negative-control comparison recovered a small amount of sensitivity at the expense of specificity (94.4% to 97.2%; 95.4% to 93.0%). Clinical notes (**Figure 6**) largely restored the specificity lost when contamination control was removed (no-contam consensus specificity 88.4% to 97.7% with notes) but did not restore the sensitivity lost when the prevalence filter was removed (91.7% with and without notes) consistent with clinical information acting as negative evidence to reject spurious detections rather than to recover pathogen signal.

### Regression testing of a taxonomic classifier update

Automated diagnosis also makes regression testing of pipeline components tractable, allowing a candidate software update to be evaluated for changes in diagnostic behaviour before adoption. As a demonstration, we compared the taxonomic classifier used throughout this study (Metabuli v1.0.5) against a development build implementing syncmer support (Metabuli v1.1.1-dev) with the reference database rebuilt accordingly (**Figure S8**). The update substantially reduced the on-disk index size (446 GB versus 875 GB; data not shown) and improved average classification speed by ∼50% (∼ 2 seconds vs ∼ 1 second; data not shown; fast classification due to small read numbers after quality control, **Table S2**). For the best performing classifier with clinical notes (*qwen3-32b-q8-tiered*) consensus diagnostic performance did not differ significantly between builds (McNemar’s test, b = 1, p = 0.5), indicating that these efficiency improvements were achieved without a detectable change in overall accuracy. Regression testing at the level of individual calls indicated a single discordant result: in a spike-in sample containing sub-threshold Murray-Valley Encephalitis virus (*Orthoflavivirus murrayense*). The model in the development build selected *Acholeplasma laidlawii* as the candidate pathogen (a mollicute commonly associated with laboratory and reagent contamination) rather than the genuine sub-threshold RNA virus. Because the selected organism did not match the spiked target, the call was scored as a false positive, reducing consensus specificity (100% to 97.7%) while leaving sensitivity unchanged.

## Supplementary Figures

**Figure S1.** Limit of detection experiment and classifier benchmark (see **Figure 1**) with a minimum RPM threshold of 10 RPM.

**Figure S2.** Limit of detection experiment and classifier benchmark (see **Figure 1**) with a minimum RPM-ratio (RPM-r) threshold (see Miller 2019) of 10 for read based classifiers used for all domains in this analysis.

**Figure S3.** Validation dataset layout with clinical samples, spike-ins and controls (n =96). Data available in **Table S1**.

**Figure S4.** Ocular fluid samples in the validation dataset (n = 22) with confirmatory testing for several positive and negative samples where metagenomic sequencing detected a pathogen or was unable to determine a pathogen (testing initiated due to lack of specific orthogonal testing information for ocular fluid samples in medical records, where only the test result was recorded but not the method). Data available in **Table S1**.

**Figure S5:** Schema of input, prompt and decision tree structures for the diagnostic classifiers (META-GPT).

**Figure S6**. Distribution of peak GPU memory and runtime for each node of the diagnostic classifiers with (blue) and without (green) clinical information for 4-bit and 8-bit quantizations (q4/q8) and decision tree architectures (single/tiered). Outliers past 10 minutes generation time are not shown (q4-tiered: n = 5, q8-tiered: n = 7, q4-single: n = 2, q8-single). Data available in **Table S5** and **Table S6**.

**Figure S7.** Reviewer and classifier replicate evaluation outcomes for the validation plate (n = 96, including controls in blue; samples below experimental limit of detection in grey). (A) Untrained reviewer outcome in the unassisted baseline evaluation without clinical notes. (B) Best performing classifier without clinical notes (qwen3-32b-q8-tiered). (C) Best performing classifier with clinical notes (qwen3-32b-q8-tiered-clinical). TP, true positive; TN, true negative; FP, false positive; FN, false negative.

**Figure S8.** Regression testing using the best performing model (qwen3-32b-q8-tiered-clinical) for a software (and associated database) update for the taxonomic classifier Metabuli, showing non-significant differences of diagnostic outcome (average sensitivity and average specificity) between the version used in this manuscript (v1.0.5) and a recent development update with syncmer support (v1.1.1-dev).

**Figure S9**: Host-taxon regressions in the Cerebro data exploration interface indicate a traceable contamination event with unknown DNA (suspected *Aspergillus flavus*) in a 0.1x TE buffer aliquot used for the external positive controls (n = 3, white points) and one of the spike-in samples (DW-63-V55, *Orthoflavivirus murrayense*, red point). We excluded several highly unusual taxonomic detections (*Aspergillus flavus*, *Sarcocystis neurona*, *Neospora caninum*, *Trypanosoma cruzi*, *Simplexvirus pteropodialpha2*, *Acholeplasma laidlawii*) that clustered at high abundance among these samples (red circle).

**Figure S10.** Quality control metrics (columns) for all DNA libraries on the validation plate (n = 96) – green indicates *pass-good*, blue indicates *pass-ok*, red indicates *failed* on the default thresholds for this study (see Methods).

**Figure S11.** Quality control metrics (columns) for all RNA libraries on the validation plate (n = 96) – green indicates *pass-good*, blue indicates *pass-ok*, red indicates *failed* on the default thresholds for this study (see Methods).

**Figure S12.** Screenshots of the WebAssembly clinical reporting interface in Cerebro (A) and the tiered interpretation interface for sample DW-63-V01 (*Streptococcus oralis*) with the optional best-species score filter disabled (B) and enabled (C).

**Figure S13.** Example of the host-taxon regression for *Toxoplasma gondii* prevalence contamination rescue, showing the two orthogonally verified samples as red dots above the Student-t prediction interval (95%) - the slope is notably positive, indicating contamination of host sequences in the reference assembly (cipher-v2-host).

**Figure S14.** Quality control metrics (columns) for all DNA libraries of the development cohort – green indicates *pass-good*, blue indicates *pass-ok*, red indicates *failed* on the default thresholds for this study (see **Methods**). Protocols were variable in this dataset.

**Figure S15.** Quality control metrics (columns) for all RNA libraries of the development cohort including failing and re-sequenced libraries (see Methods) – green indicates *pass-good*, blue indicates *pass-ok*, red indicates *failed* on the default thresholds for this study (see **Methods**). Protocols were variable in this dataset.

## Supplementary Tables

**Table S1.** Validation dataset reference testing and review results. Patient scenarios in the table for the spike in studies are fabricated for the purposes of the study

**Table S2.** Validation dataset sequencing results and quality control.

**Table S3.** Validation dataset taxa and classifier evidence after stratification and filtering.

**Table S4.** Validation dataset memory usage for diagnostic classifier configurations and replicates.

**Table S5.** Validation dataset runtime for each prompt in the decision tress for diagnostic classifier configurations and replicates.

**Table S6.** Development cohort results for expert review and diagnostic classifiers.

**Table S7.** Development cohort sequencing results and quality control.

**Table S8.** Development cohort taxa and classifier evidence after stratification and filtering.

## Notes

### Competing Interest Statement

The authors have declared no competing interest.

### Author Declarations

Ethics committee of Melbourne Health gave ethical approval for this work: (HREC 74254)

