## Supplementary figures and images for "Large language models enable consensus-level interpretation in metagenomic diagnostics"

### Figure_S1.png

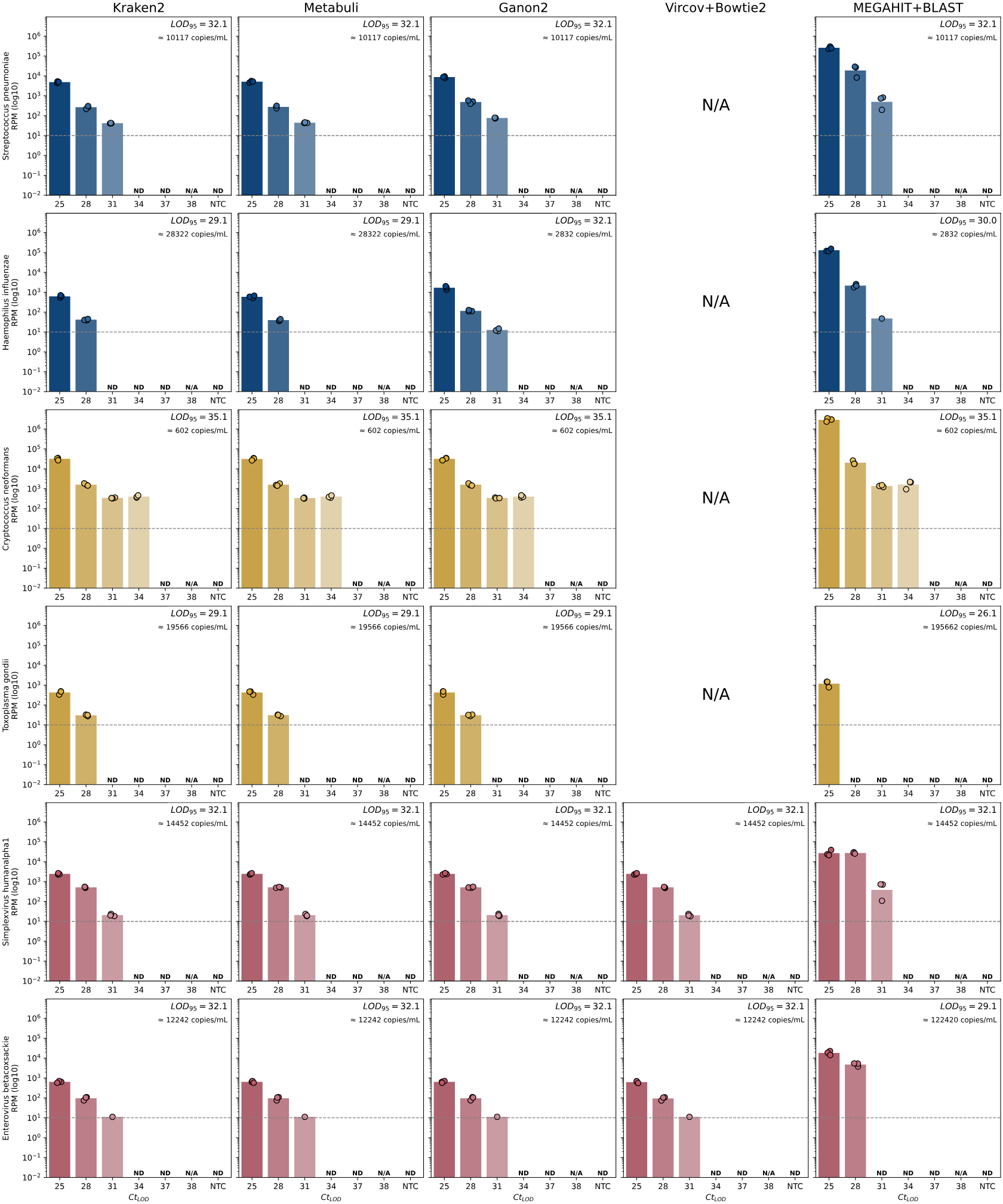

### Figure_S2.png

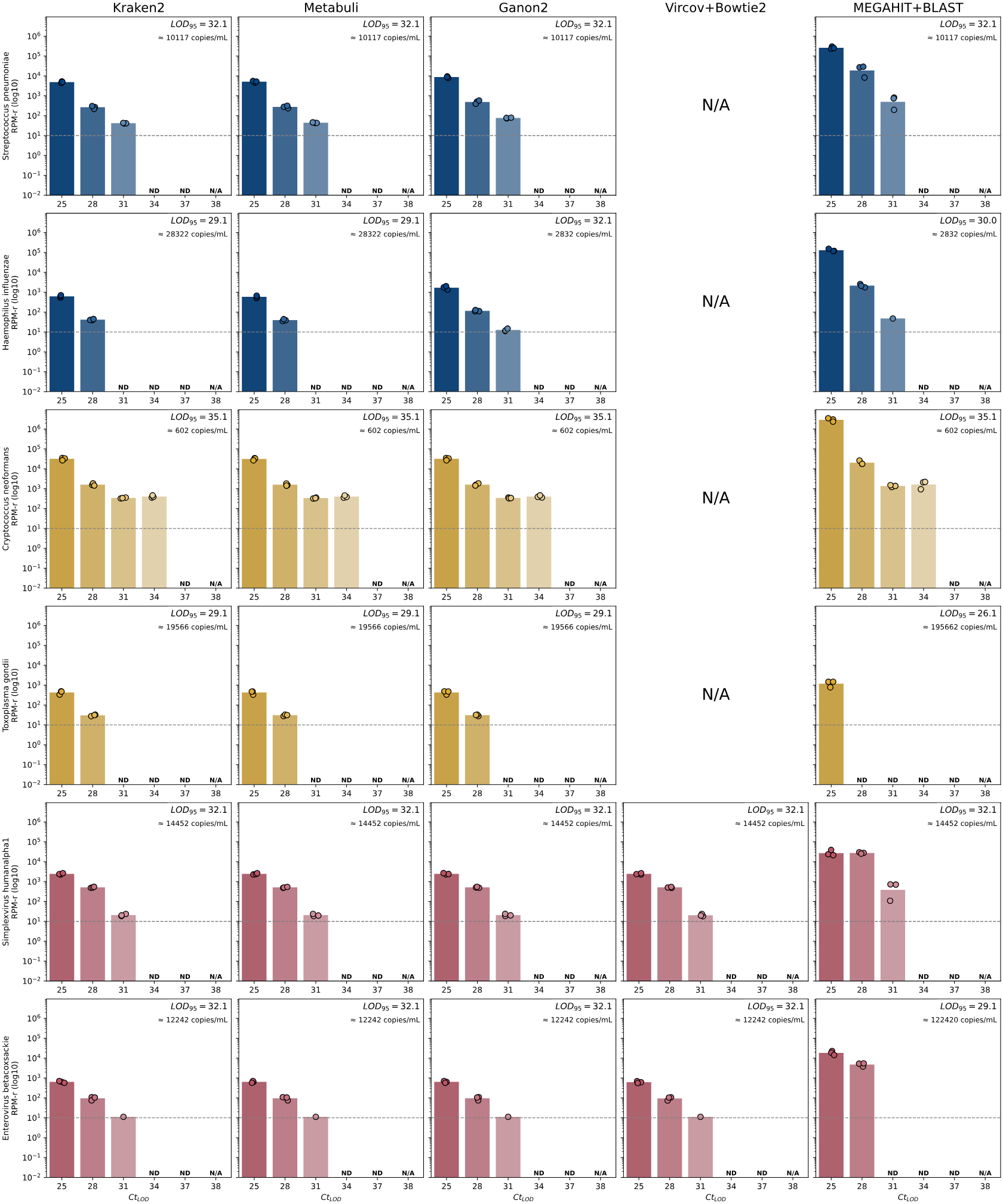

### Figure_S3.png

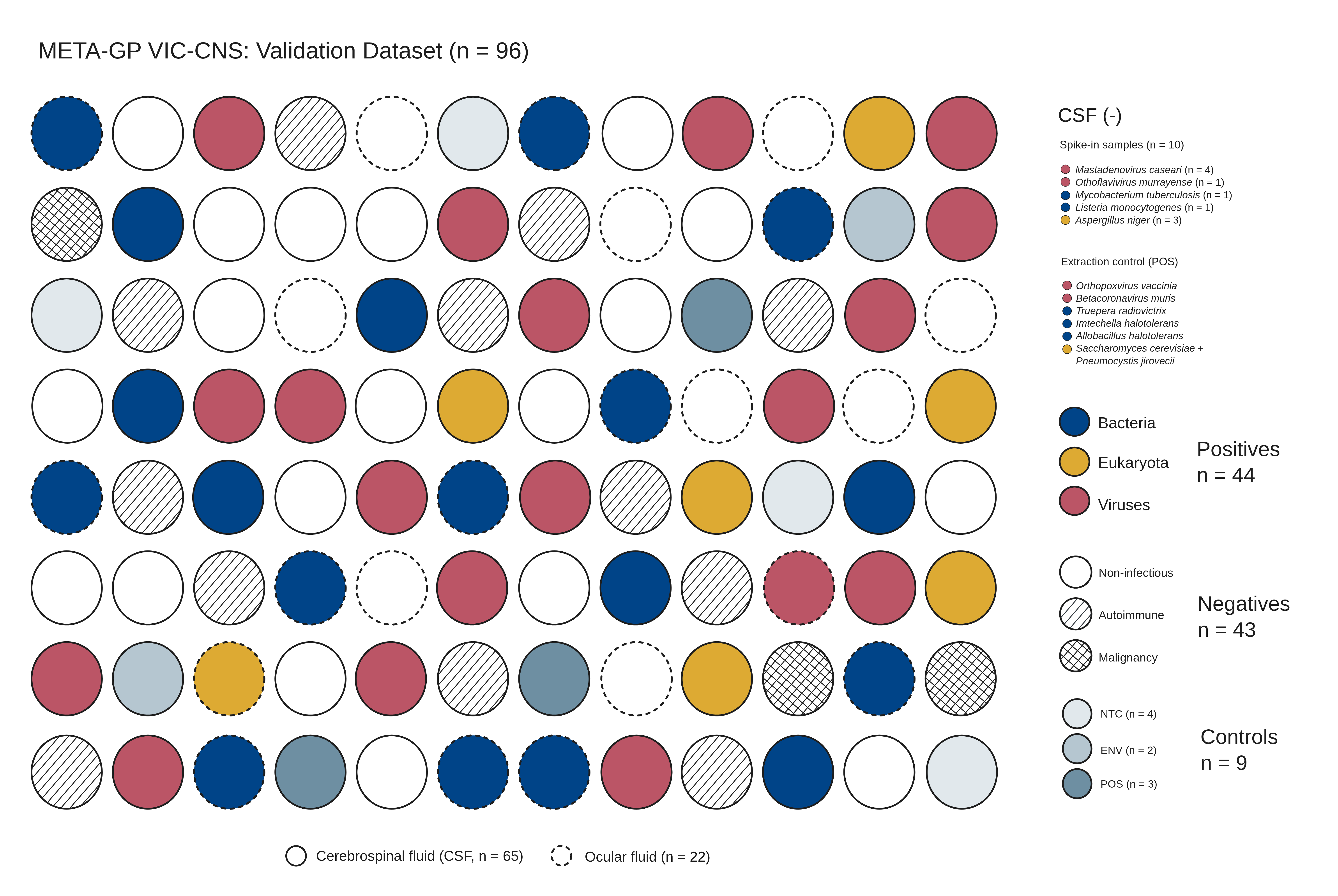

### Figure_S4.png

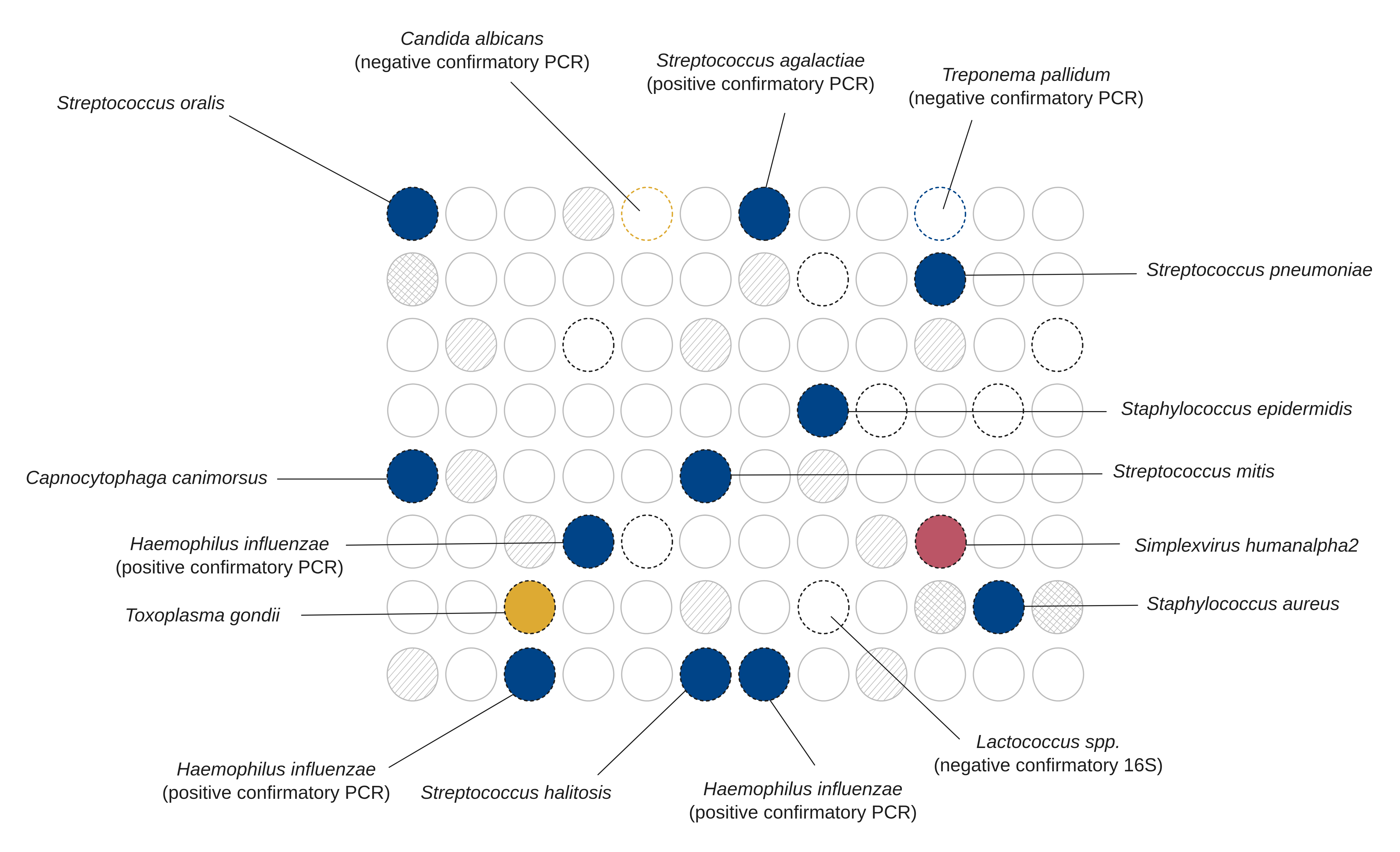

### Figure_S5.png

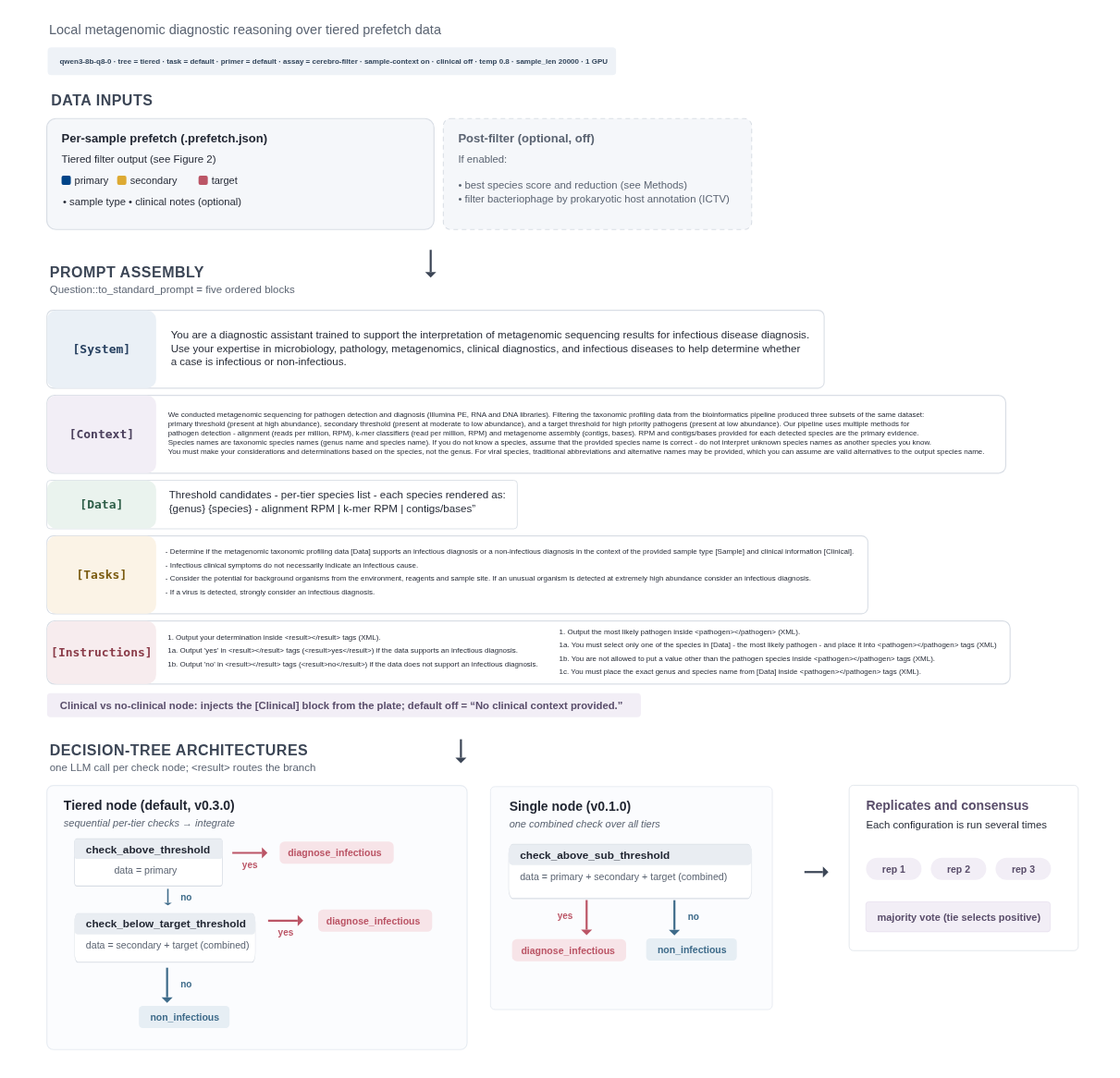

### Figure_S6.pdf

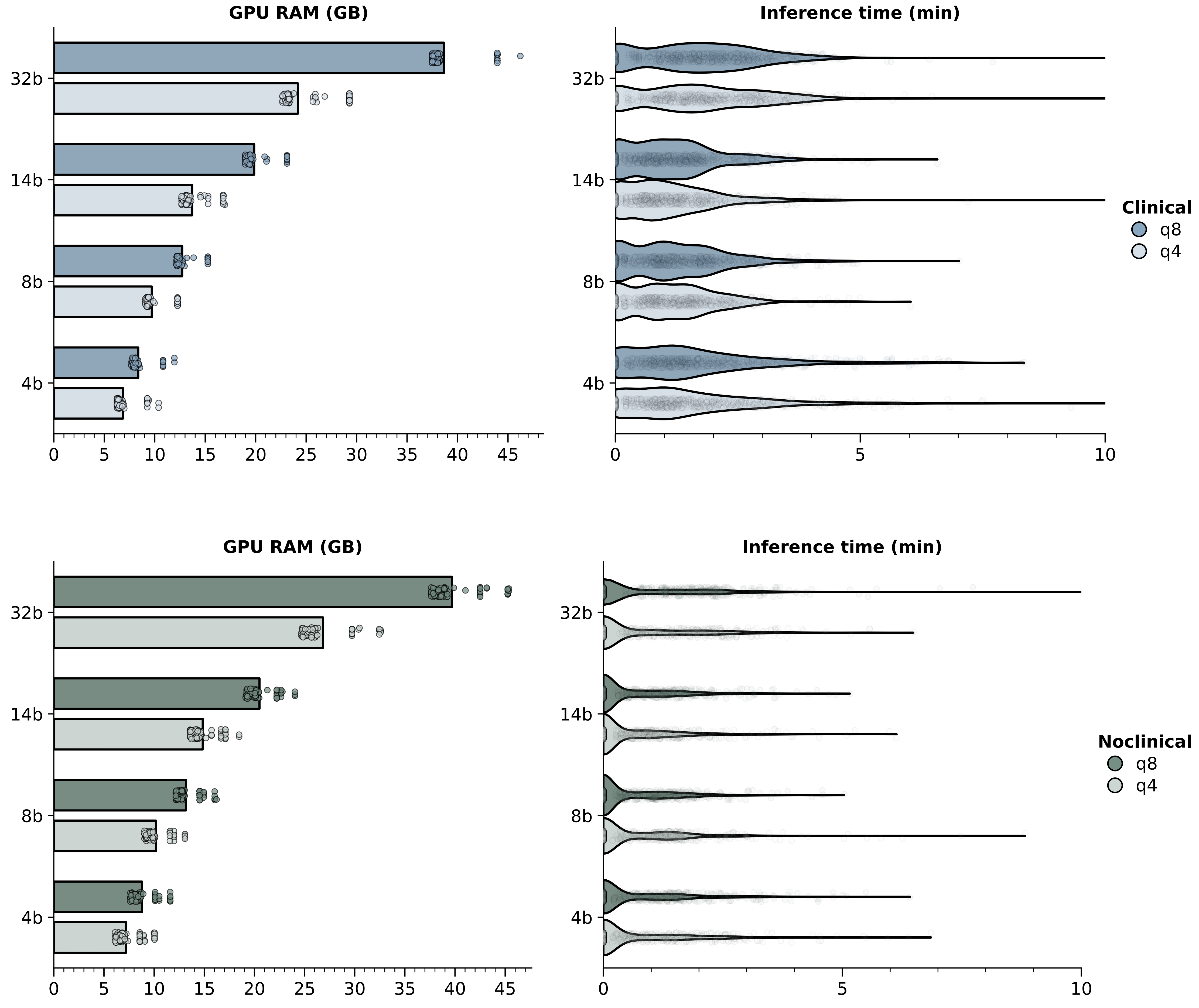

### Figure_S7.png

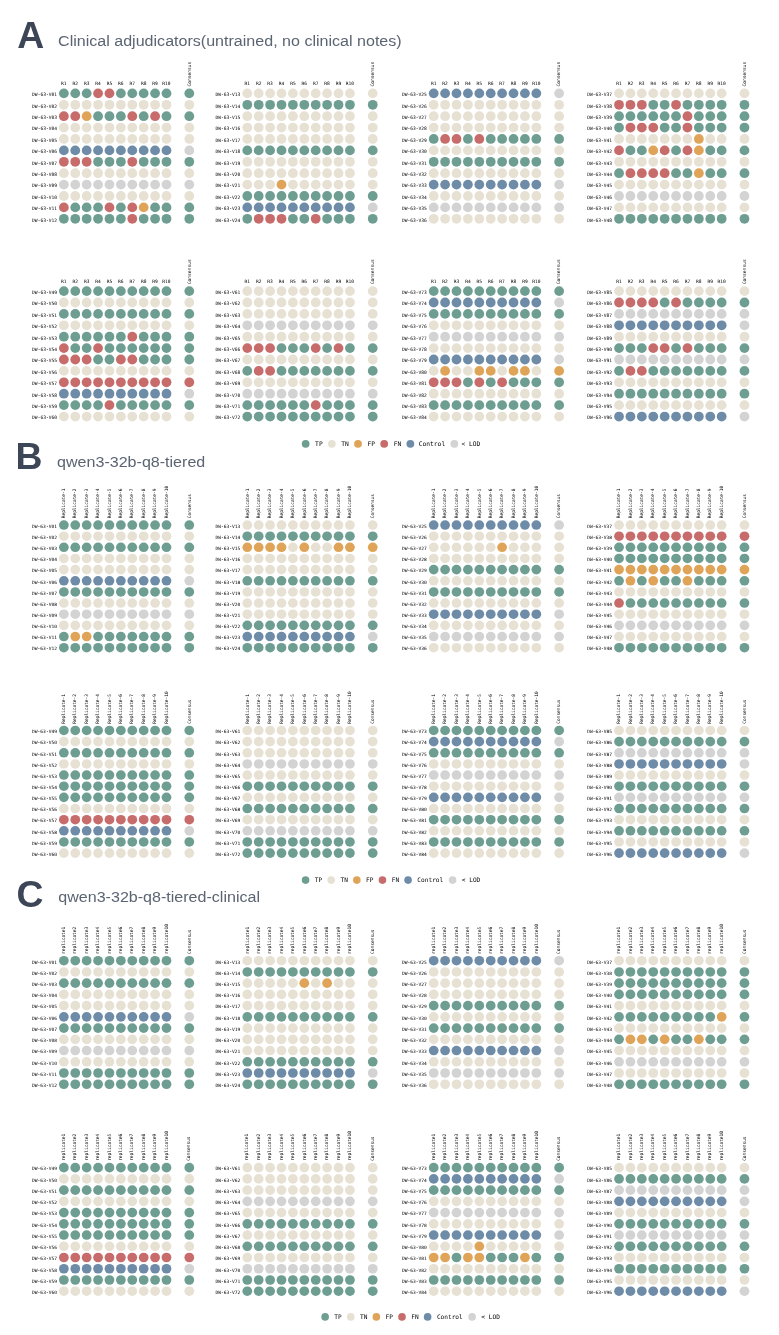

### Figure_S8.png

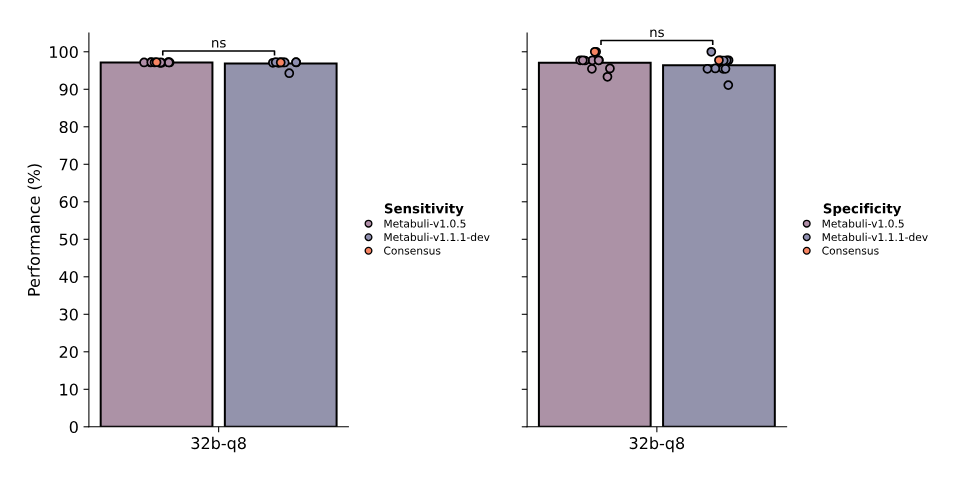

### Figure_S9.png

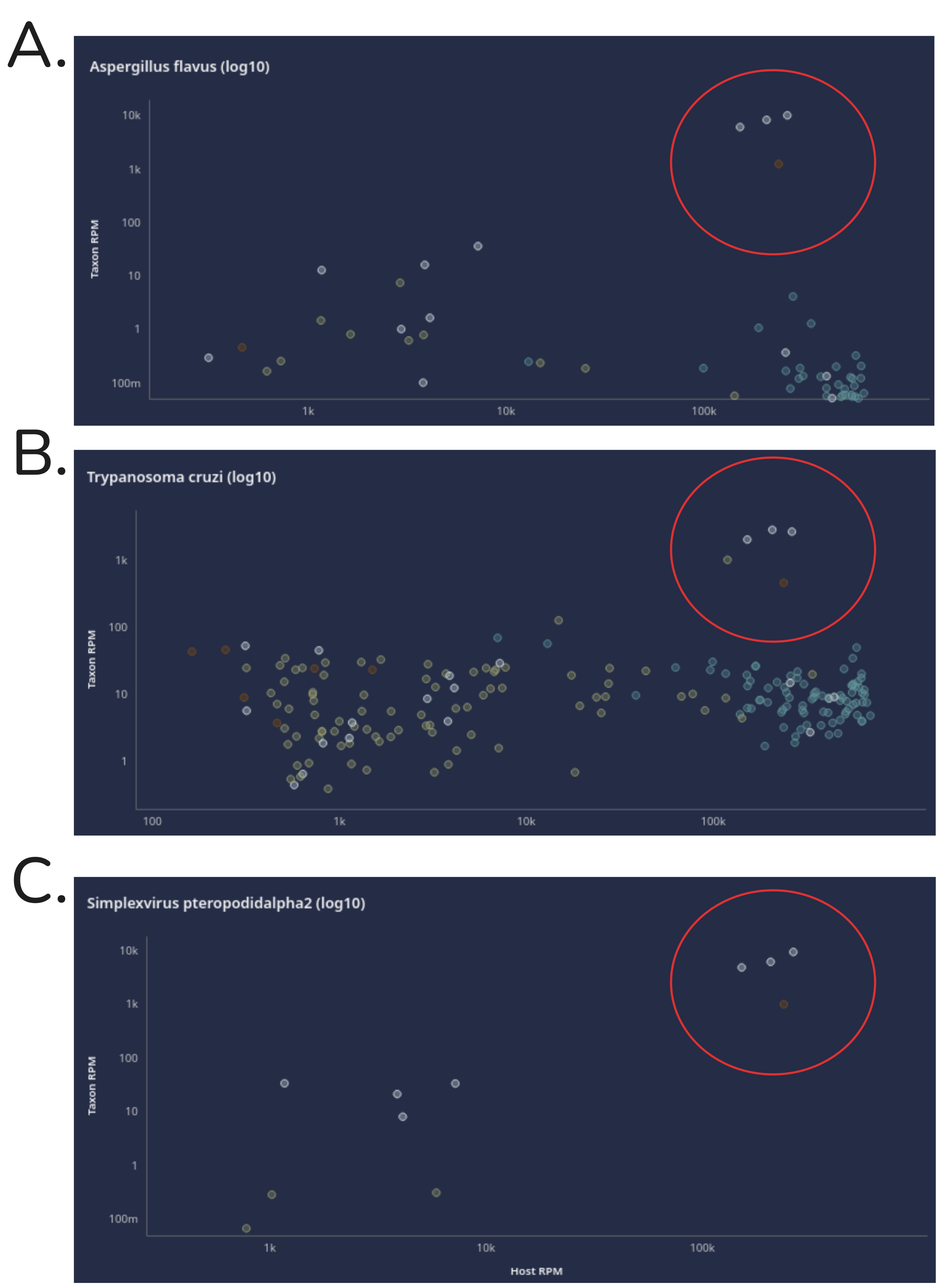

### Figure_S10.png

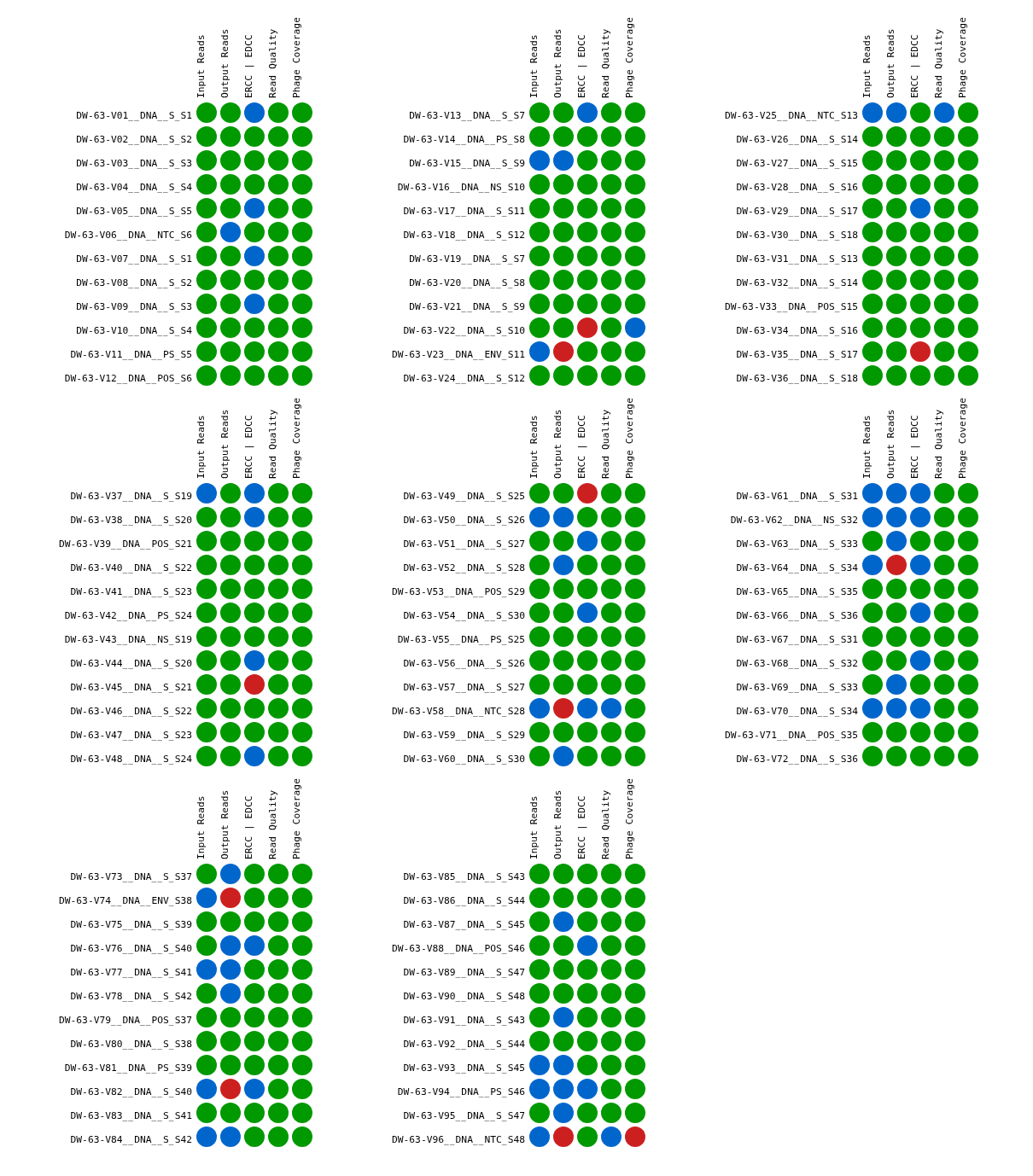

### Figure_S11.png

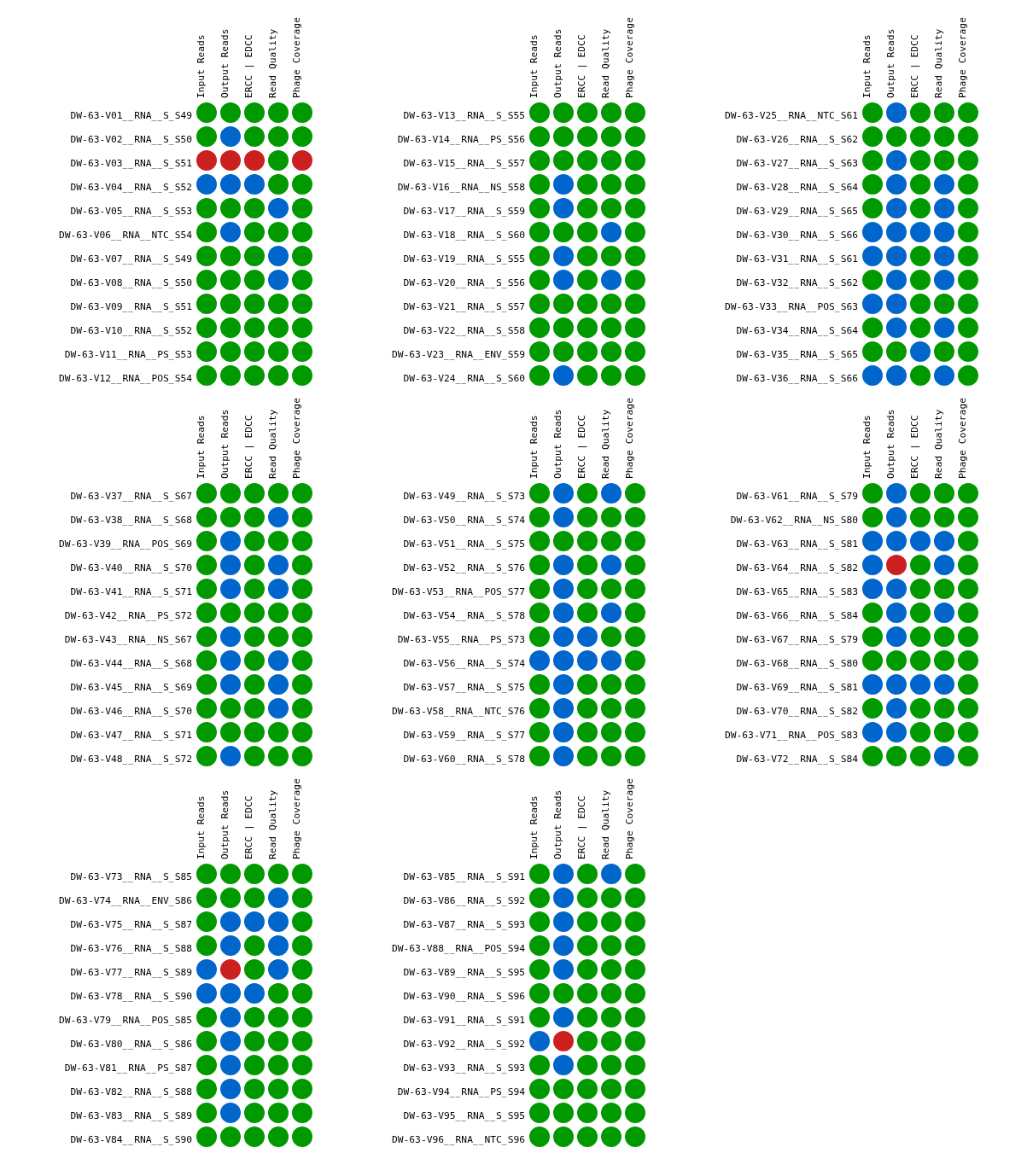

### Figure_S12.png

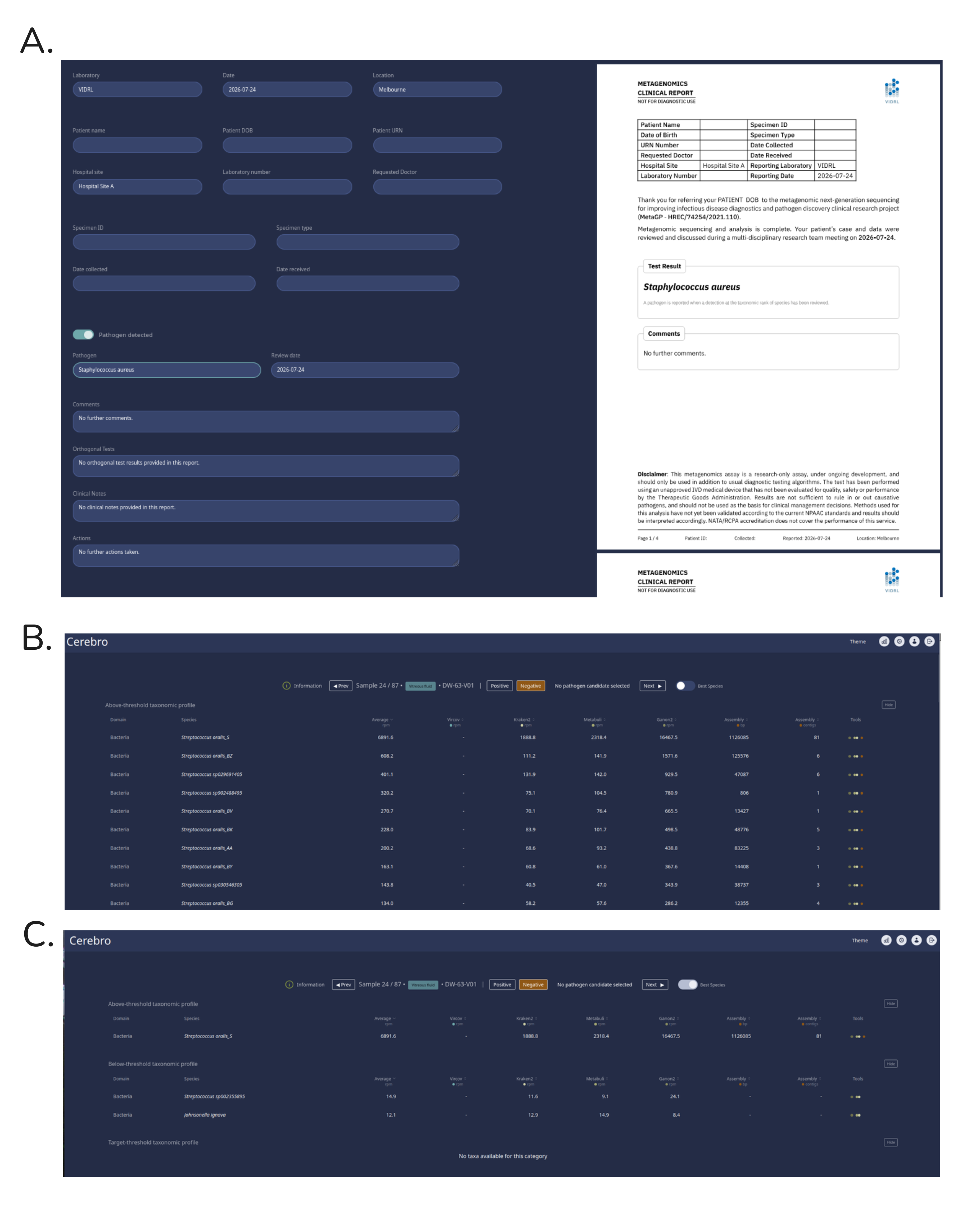

### Figure_S13.png

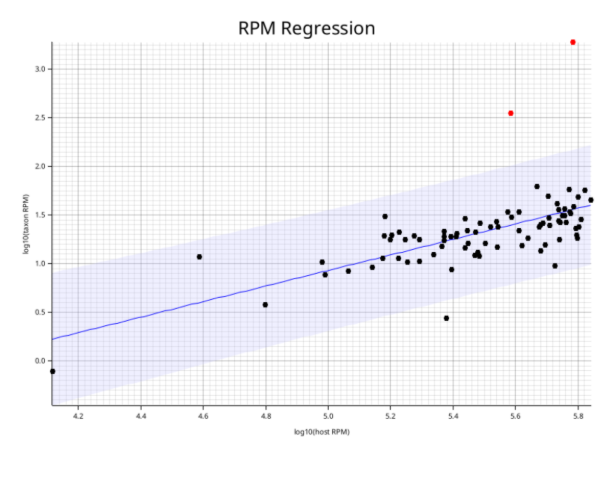

### Figure_S14.png

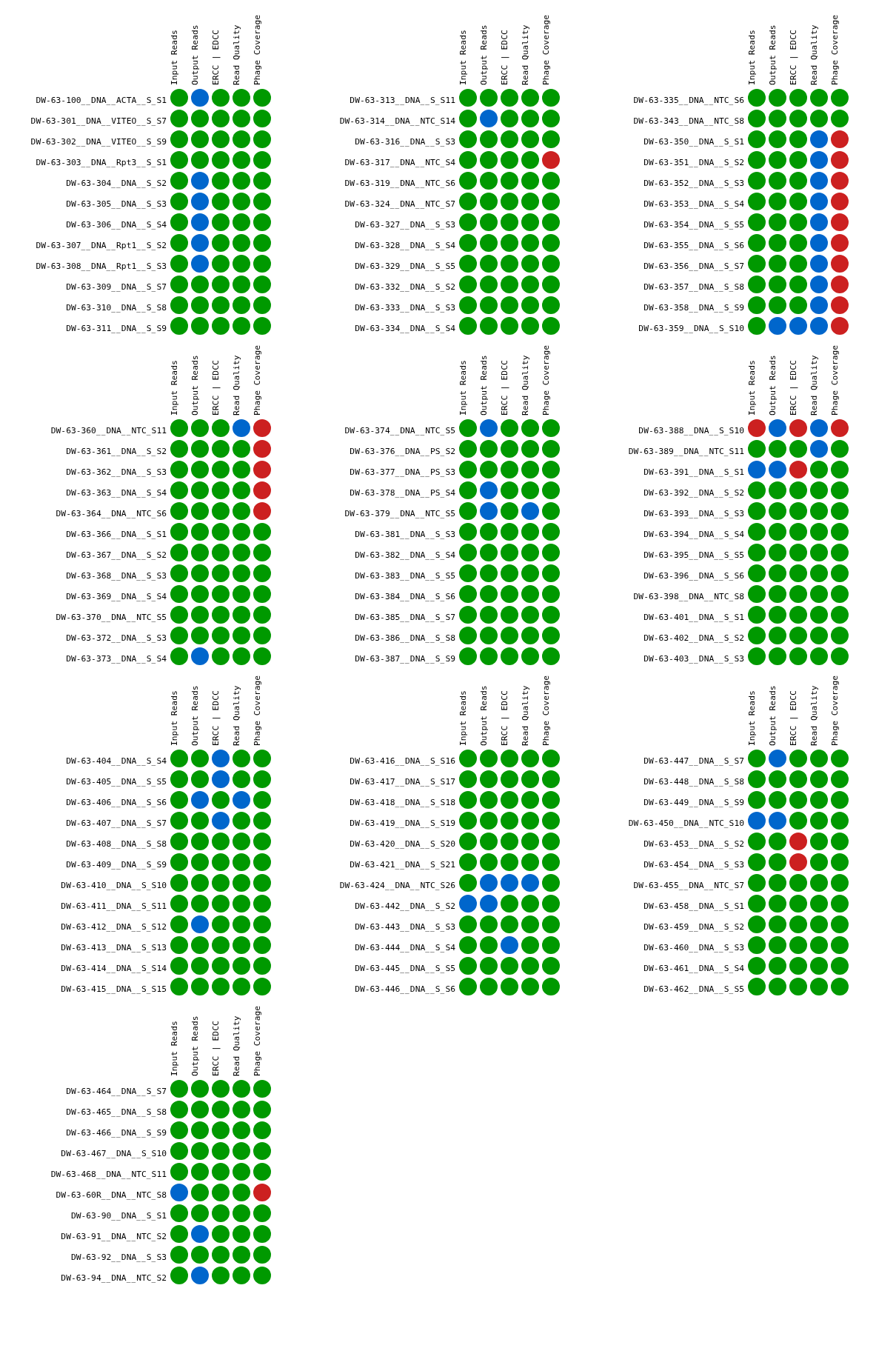

### Figure_S15.png

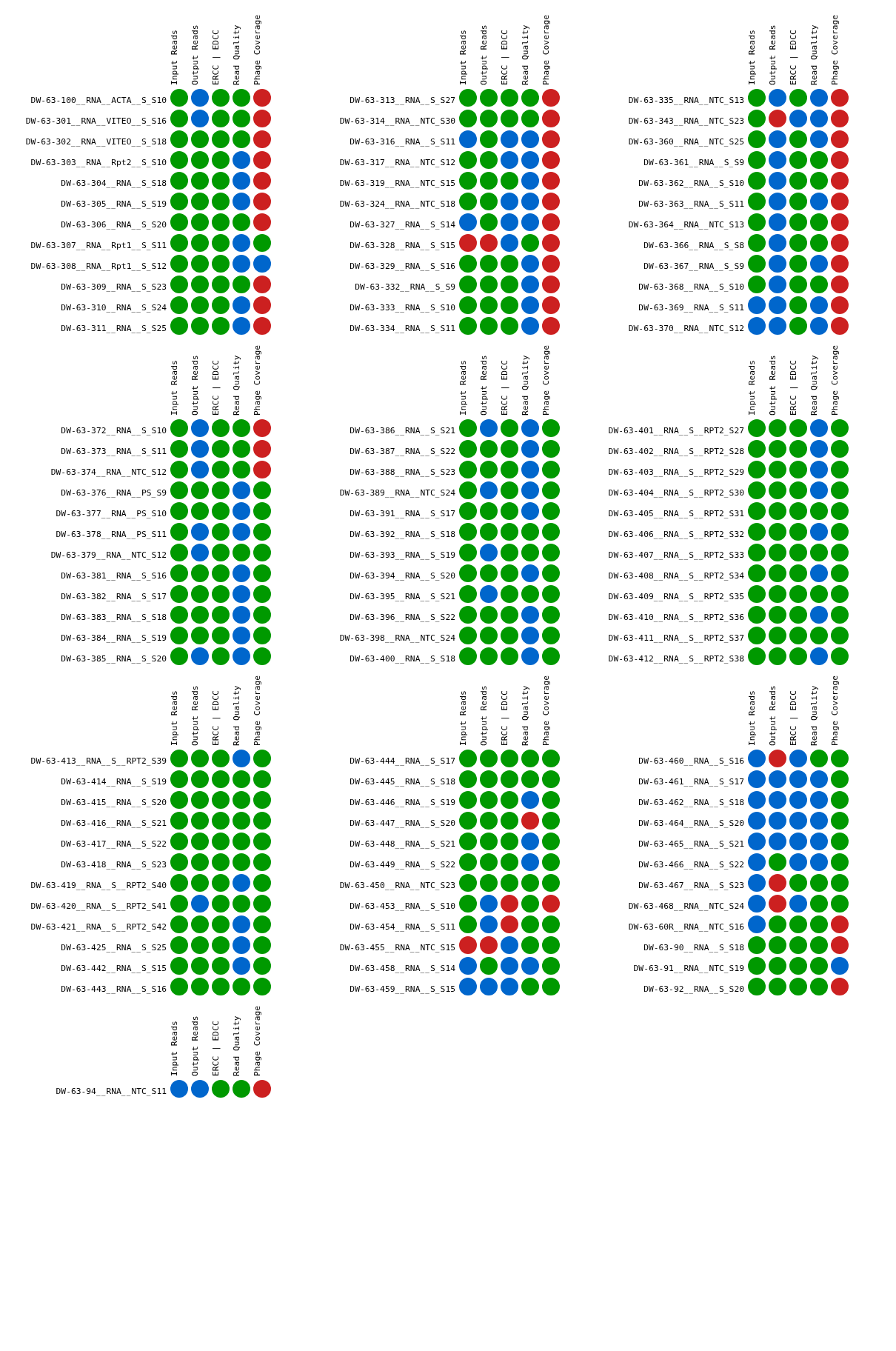
